# Biofabrication of multiplexed electrochemical immunosensors for simultaneous detection of clinical biomarkers in complex fluids

**DOI:** 10.1101/2022.03.18.22272576

**Authors:** Sanjay S. Timilsina, Mohanraj Ramasamy, Nolan Durr, Rushdy Ahmad, Pawan Jolly, Donald E. Ingber

## Abstract

Simultaneous detection of multiple disease biomarkers in unprocessed whole blood is considered the gold standard for accurate clinical diagnosis. Here, we report the development of a 4-plex electrochemical (EC) immunosensor with on-chip negative control capable of detecting a range of biomarkers in small volumes (15 µL) of complex biological fluids, including serum, plasma, and whole blood. A framework for fabricating and optimizing multiplexed sandwich immunoassays is presented that is enabled by use of EC sensor chips coated with an ultra-selective, antifouling, nanocomposite coating. Cyclic voltammetry evaluation of sensor performance was carried out by monitoring the local precipitation of an electroactive product generated by horseradish peroxidase linked to a secondary antibody. EC immunosensors demonstrated high sensitivity and specificity without background signal with a limit of detection in single-digit pg/mL in multiple complex biological fluids. These multiplexed immunosensors enabled simultaneous detect of four different biomarkers in plasma and whole blood with excellent sensitivity and selectivity. This rapid and cost-effective biosensor platform can be further adapted for use with different high affinity probes for any biomarker, and thereby create for a new class of highly sensitive and specific multiplexed diagnostics.

## 1. Introduction

The ongoing paradigm shift from reactive healthcare to preventive health care has increased interest in patient-centric diagnostic devices that could be used in everyday life. Direct detection of relevant disease biomarkers in whole blood could circumvent expensive and time-consuming sample preparation procedures that are currently required in clinical laboratories so that diagnotic assays could be performed in point-of-care (POC) settings, including physicians offices, pharmacies, and even at home.^[1]^ Widely used detection techniques like optical detection cannot be used for whole blood detection because of strong scattering, absorption, and considerable autofluorescence in these complex biological samples.^[2]^ Complex biological fluids, such as whole blood, plasma, and serum are also difficult to analyze due to high concentrations of sticky proteins and other molecules that can reduce signal detection and increase background noise. While some of these negative effects can sometimes be partly reduced by sample dilution, many biomarkers are present near the assay’s detection limit, and the dilution of samples is not always linear.^[3]^

One potential way to overcome these diagnostic challenges is through the use of electrochemical (EC) sensors that can provide rapid, accurate, and quantitative detection of disease biomarkers in complex fluids.^[4]^ Because they generate an electrical signal as an output, they also can seamlessly integrate with wireless data collection and transmission systems. But while significant basic research has been conducted on developing multiplexed EC POC devices for disease diagnosis, very few products have been translated to the clinical setting.^[5]^ There are various reasons for this, including biofouling of electrodes by components with complex biological fluids (sweat, saliva, blood, plasma), the complexity of building bioassays on the electrodes, challenges related to development of multiplexing capabilities, sensitivity to changes in environmental cues, and the need to develop sophisticated readouts and analytical tools for analysis of complex data generated with these types of devices.^[6]^

We have recently developed an engineering solution to address these challenges and create a clinically translatable EC platform for rapid detection and quantification of clinically relevant disease biomarkers in small volumes of complex biological fluids, including whole human blood. Key to this advance was the development of a novel antifouling electroconductive nanocomposite coating to combat EC biosensors, which enabled engineering of an EC sensor platform that provides effective and accurate detection of several biomarkers that may be used for diagnosis of various diseases with clinical significance.^[5, 7]^ The coating method involves brief (1 minute), localized, heat-induced coating of EC sensors with a nanocomposite composed of denatured bovine serum albumin cross-linked with pentaamine functionalized graphene oxide cross-linked with glutaraldehyde (BSA/prGOx/GA).^[8]^

Multiplexing is a critical requirement for POC devices because disease and pathophysiology often involves interplay among many complex biological processes, and hence accurate diagnosis requires detection of several molecules rather than a single biomarker entity.^[4c, 9]^ Establishment of a multiplexed detection platform therefore makes it possible to stratify and monitor complex multifactorial diseases with high confidence in conjunction with relevant validated biomarkers.^[10]^ In addition, multiplexing minimizes assay costs, time, and sample volume while concurrently enabling efficient monitoring and prediction of disease progression and outcome.^[11]^ To develop this type of multiplexed platform, it is imperative to immobilize optimized concentrations of high affinity bioreceptor molecules for the target of interest on the electrode surface, which in combination with a specific detection antibody provides a high level of signal sensitivity. The coating density of bioreceptors on the electrode surface is critical in achieving the optimal surface-to-volume ratio necessary for the efficient capture and detection of the biomarker in the test sample.^[12]^

Here, we describe methods and tools that can be used to biofunctionalize nanocomposite coated multi-electrode sensor chips and to assess sensor functionality to detect multiple analytes in the presence of complex biological fluids. A sandwich enzyme-linked immunosorbent assay (ELISA) is employed for signal detection that uses secondary antibodies linked to horseradish peroxidase (HRP) enzyme, which generate an electroactive, insoluble, 3,3,5,5-Tetramethylbenzidine (TMB) product that precipitates locally at the molecular binding site above the electrode surface, and thereby enables a multiplexing capability. Different assay parameters were optimized to create an immunoassay with optimum probe density and TMB precipitation to develop a multiplexed assay with required sensitivity and specificity without electrochemical background signal. Using this approach, an immunoassay with single-digit pg/mL sensitivity was developed that was not influenced by clinically relevant hematocrit levels in whole blood, which can potentially be translated into development of diagnostics for POC settings. We describe how we used framework for sensor assay development to create optimized EC sensors that enable multiplexed detection of a range of protein biomarkers for multiple clinically relevant conditions and disease, including sepsis, active pulmonary tuberculosis (TB), myocardial infarction (MI), traumatic brain injury (TBI), and multiple sclerosis (MS).

## 2. Results

### 2.1 Fabrication of electrochemical biosensors

The methods used for fabrication of our EC sensors are critical for their high functionality. Clean gold sensor electrodes were first treated with plasma cleaner for 8 min (**Figure S1**, Supporting Information) and modified with bioinspired nanocomposite coating composed of glutaraldehyde (GA) cross-linked with denatured bovine serum albumin (BSA) intercalated with pentaamine modified reduced graphene nanoparticles (prGOx) via rapid coating method as described previously^[8a]^ prior to immobilizing antibodies (**Figure 1**). Importantly, we also found that this antifouling coating application method can also be applied to other relevant electrode materials, such as a gold electroplated printed circuit board (PCB) on polyethylene terephthalate (PET) substrate from Linxens and a 3D graphene foam-based sensing electrode (Gii-Sens, from Integrated Graphene). Cyclic voltammetry (CV) analysis of the coated Linxens sensors confirmed that they maintain similar currents as the bare electrode (**Figure S2**, Supporting Information). On the other hand, a slight decrease in the peak current was observed after coating the Gii-sens electrodes, which could be attributed to the porous structure of graphene foam, which was filled after applying the coating (**Figure S3a**, Supporting Information).

**Figure 1:**
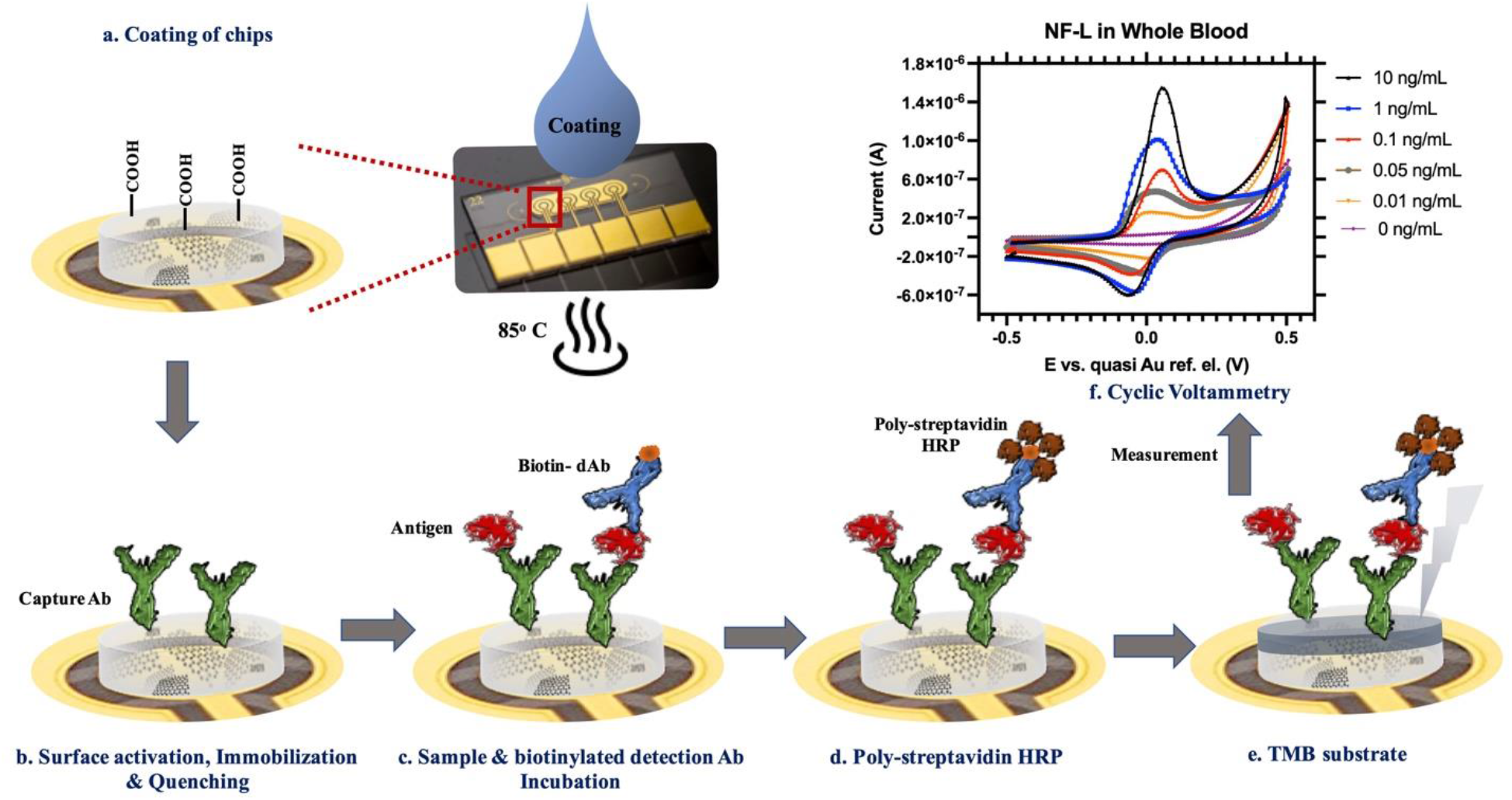
3D schematic of sandwich immunoassay on biosensor coated with antifouling nanocomposite. a) Electrochemical sensor with gold electrodes coated with the antifouling nanocomposite at 85 °C for 45s. b) Immobilization of capture antibody followed by quenching and blocking with bovine serum albumin. c) Incubation of sample mixed with biotinylated detection antibodies. d) Addition of streptavidin poly-HRP. e) Addition of precipitating TMB and subsequent precipitation over the gold electrode in EC biosensor. f) Cyclic voltammogram for the measurement of NF-L in whole blood using biosensor with anti-fouling coating.

The coating on these sensors was then activated with 1-ethyl-3-[3-dimethylami-nopropyl]-carbodiimide hydrochloride/ N-hydroxysuccinimide (EDC/NHS) and incubated with bioreceptor molecules (i.e., capture antibodies), resulting in the formation of covalent links between free carboxylate groups of the coating and primary amines of the antibody; unreacted carboxylate groups were subsequently quenched with ethanolamine (**Figure 1b**). Although there was a slight decrease in the current with coated Gii-sens electrodes, functionalization of the coated electrode directly with HRP enzyme (as a positive control) only showed a high signal when its TMB substrate was added with no signal in negative control (**Figure S3b**, Supporting Information). Thus, these findings indicate that the antifouling nanocomposite coating can be applied to various kinds of sensors and support selective binding of desired capture molecules on top of the electrode surface.

Coated immunosensors with immobilized capture antibodies were then exposed to samples containing the antigen and a secondary biotinylated detection antibody. Upon incubation, the specific antigen is sandwiched between the capture and biotinylated detection antibodies (**Figure 1c**). Next, the sensors are washed and exposed to streptavidin poly-horseradish peroxidase (spHRP), which binds to the biotinylated detection antibody (**Figure 1d**). Finally, the TMB substrate for the HRP enzyme is added, which results in production of an insoluble electroconductive product that locally precipitates at the reaction site and is electrochemically read using cyclic voltammetry (**Figure 1e**). Typical CVs are obtained with the immunosensor, where the peak height is directly proportional to the amount of target detected (**Figure 1f**).

### 2.2 Electrochemical immunoassay development

The nanocomposite-coated EC sensors can be used to develop both two-step (incubation of sample followed by washing and addition of detection antibody) and single-step (incubation with pre-mixed sample and detection antibody) assays; however, their effectiveness requires optimization of critical reagents and processes that are used to define assay performance. Using the nanocomposite coating, we found that the highest sensitivity and specificity were achieved by optimizing the concentration of capture and detection antibody, spHRP, as well as the TMB incubation time. Initially, a two-step assay was evaluated using the MI biomarkers, cardiac troponin I (cTnI) and N-terminal (NT)-pro hormone BNP (NT-proBNP), as targets. These biomarkers show variable levels in circulation according to the clinical condition and timing of measurement and thus require a rapid and sensitive POC device for clinical use.^[13]^ Plasma samples were added to the EC sensor followed by an optimized concentration of 1 μg/mL detection antibody to achieve a sensitivity of 24 and 3 pg/mL for cTnI and NT-proBNP, respectively, with assay times of 1 h and 21 min (**Figure S4**, Supporting Information). The EC sensor also was not able to achieve the clinical threshold for cTnI, which is 16 pg/ml in female.^[13b]^

Assay development was then carried out to develop a rapid biosensor with decreased assay time while maintaining high sensitivity and specificity for single-step detection of the different biomarkers. Surface coverage plays a critical role in defining the performance of the EC sensor and can be optimized to reduce steric hindrance for efficient binding. Different concentrations of anti-cTnI capture antibody (50 µg/mL-1000 µg/mL) were evaluated to detect high, low and zero concentrations of the analyte (10, 0.1, and 0 ng/mL, respectively). The electrical signal generated with the 10 ng/mL analyte increased as the concentration of the capture antibody was raised from 50 to 500 µg/mL as expected but it decreased when the antibody concentration was further raised from 500 to 1000 µg/mL, which may be due to surface crowding (**Figure 2a**).^[14]^ 500 µg/mL of capture antibody also resulted in the highest signal-to-noise ratio, and a significant difference between lowest concentration and blank. This 500 µg/mL capture antibody concentration was then used to optimize the detection antibody concentration in the range of 1-8 µg/mL with high, low, and blank analyte.

**Figure 2:**
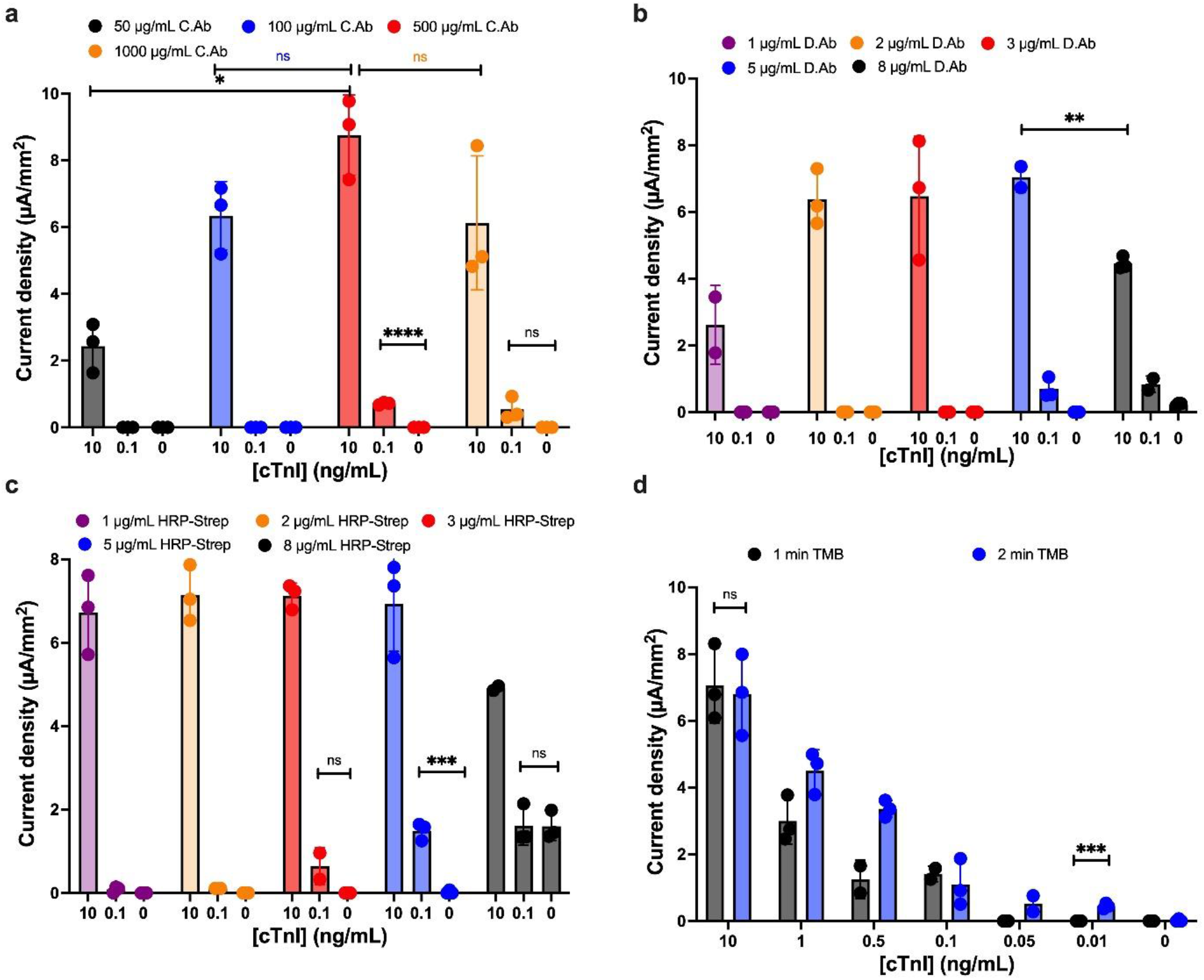
Optimization of assay condition for detection of cTnI in EC biosensor. a) Optimization of cTnI capture antibody. Bar graph shows the mean current density for different concentration of capture antibody (50, 100, 500, and 1000 μg/mL) to perform assay of cTnI at 3 different concentrations (10, 0.1, and 0 ng/mL). b) Optimization of cTnI detection antibody. Bar graph shows the mean current density for different concentration of detection antibody (1, 2, 3, 5, and 8 μg/mL) to perform assay of cTnI at 3 different concentrations (10, 0.1, and 0 ng/mL). c) Optimization of Streptavidin-poly-HRP (spHRP). Bar graph shows the mean current density for different concentration of spHRP (1, 2, 3, 5, and 8 μg/mL) to perform assay of cTnI at 3 different concentrations (10, 0.1, and 0 ng/mL). d) Optimization of TMB incubation time. Bar graph shows the mean current density for different incubation time for TMB (1 and 2 min) to perform assay of cTnI at different concentrations (10, 1, 0.5, 0.1, 0.05, 0.01, and 0 ng/mL). Error bars represent the s.d. of the mean; n=3. Significant difference was determined by unpaired two-tailed t-test (ns P > 0.05; *P<0.05; **P<0.01; ***P<0.001; ****P<0.0001).

Using this optimized biosensor, we found that the signal for 10 ng/mL analyte increased when the concentration of anti-cTnI detection antibody was raised from 1 to 2 µg/mL, the signal then plateaued from when the concentration was further increased from 2 to 5 µg/mL, and it finally declined from 5 to 8 µg/mL (**Figure 2b)**, which could be due to over precipitation of TMB making the surface less conductive. A signal for lower concentration (0.1 ng/mL) of analyte was seen only for detection antibody concentrations of 5 µg/mL and above, while 8 µg/mL of detection antibody resulted in non-specific binding similar to that observed for the blank.

We then used 5 µg/mL of detection antibody that had the highest signal-to-noise ratio to further optimize spHRP. Using 10 ng/mL of analyte, we found the signal generated to be consistent when we added 1 to 5 µg/mL spHRP, but with higher concentrations, there was over precipitation of substrate leading to a decrease in signal (**Figure 2c)**. Lowest concentration (0.1 ng/mL), started showing signal when concentration of spHRP was 3 µg/mL or higher with no significant difference between lowest concentration and blank at 3 µg/mL of spHRP. At 5 µg/mL of spHRP, there was a significant difference between 0.1 ng/ml and blank. However, a high non-specific signal was observed at 8 µg/mL spHRP. Thus, we viewed 5 µg/mL spHRP to be the optimal concentration for use in these studies.

Finally, all the optimized conditions were used to evaluate the TMB precipitation time. Calibration curves were run from 0.01 to 10 ng/mL with a TMB precipitation time of 1 versus 2 min. With the 1 min precipitation time, no signal was observed for lower TMB concentrations (0.05 and 0.01 ng/mL); however, signals were observed over the whole calibration range with the 2 min precipitation (**Figure 2d**). Longer precipitation times led to a non-specific signal, so the 2 min TMB precipitation time was considered optimum for the assay.

The coating time of the sensor with antifouling nanocomposite was also assessed by performing the EC sandwich assay for detection of 4 different concentrations (1, 0.1, 0.05, and 0 ng/mL) of cardiac troponin complex (cTnITC). We found that there was no significant difference at higher concentrations, but the 45 sec coating time gave the highest signal-to-noise ratio at lower analyte concentration (0.05 ng/mL) (**Figure S5a**; Supporting Information), and thus coating time of 45 sec was used for all susequent studies. Interestingly, we found that this 45 sec rapid coating procedure resulted in production of bioassays that were as sensitive as those previously created using a 24 h coating^[8b]^, as demonstrated by the absence of any significant difference in the signal generated in the cTnITC assay at lower analyte concentrations (**Figure S5b**, Supporting Information). In addition, the biosensors can be stored at room temperature for at least a week after the completion of the assay^[8a]^ and still generate a similar signal. For example, when we ran a full calibration curve (0.01 to 10 ng/mL cTnITC) there was no significant difference between the signal read out immediately after sensor fabrication or after incubating in the dark for 24 h (**Figure S5c**, Supporting Information).

As the ultimate aim was to perform a multiplexed assay with the same reagents, the spHRP concentration and TMB precipitation time were kept constant, and only the detection antibody concentration was optimized for the other biomarkers. Similar to optimization of the anti-cTnI detection antibody, we found that 9 µg/mL of anti-BNP detection antibody and 6 µg/mL of anti-NT-proBNP and anti-cTnITC complex detection antibodies gave the highest signal-to-noise ratios (**Figure S6a-c**, Supporting Information). Furthermore, similar to the biomarkers for MI, we also optimized capture and detection antibody concentration for previous described biomarkers for active pulmonary TB.^[15]^. We found that 1 mg/mL was the optimal capture antibody concentration for interleukin-6 (IL-6), IL-8, IL-18, and vascular endothelial growth factor (VEGF) using an optimal concentration of detection antibody of 2 µg/mL (**Figure S7**,**8**, Supporting Information).

### 2.2 EC biosensor analytical performance

To explore the potential value of this approach for clinical diagnostics, we leveraged these biomarkers to construct a prototype POC affinity-based ELISA on miniaturized electrodes, which could be used to develop a low-cost, rapid, ultra-low volume, and easily accessible blood-based triage test for TB. We performed sandwich ELISA on the EC biosensor containing the immobilized capture antibodies for the TB biomarkers described above and carried out calibration curves covering their clinical ranges **(Figure 3)**. We obtained a limit of detection (LOD) of 1000, 3, 9, 5, and 17 pg/mL for Ag85-B, IL-6, IL-8, IL-18, and VEGF, respectively. Importantly, the LODs for all of the TB biomarkers were better than the clinical cut-off values (**Table 1**, Supporting Information).^[15-16]^

**Figure 3:**
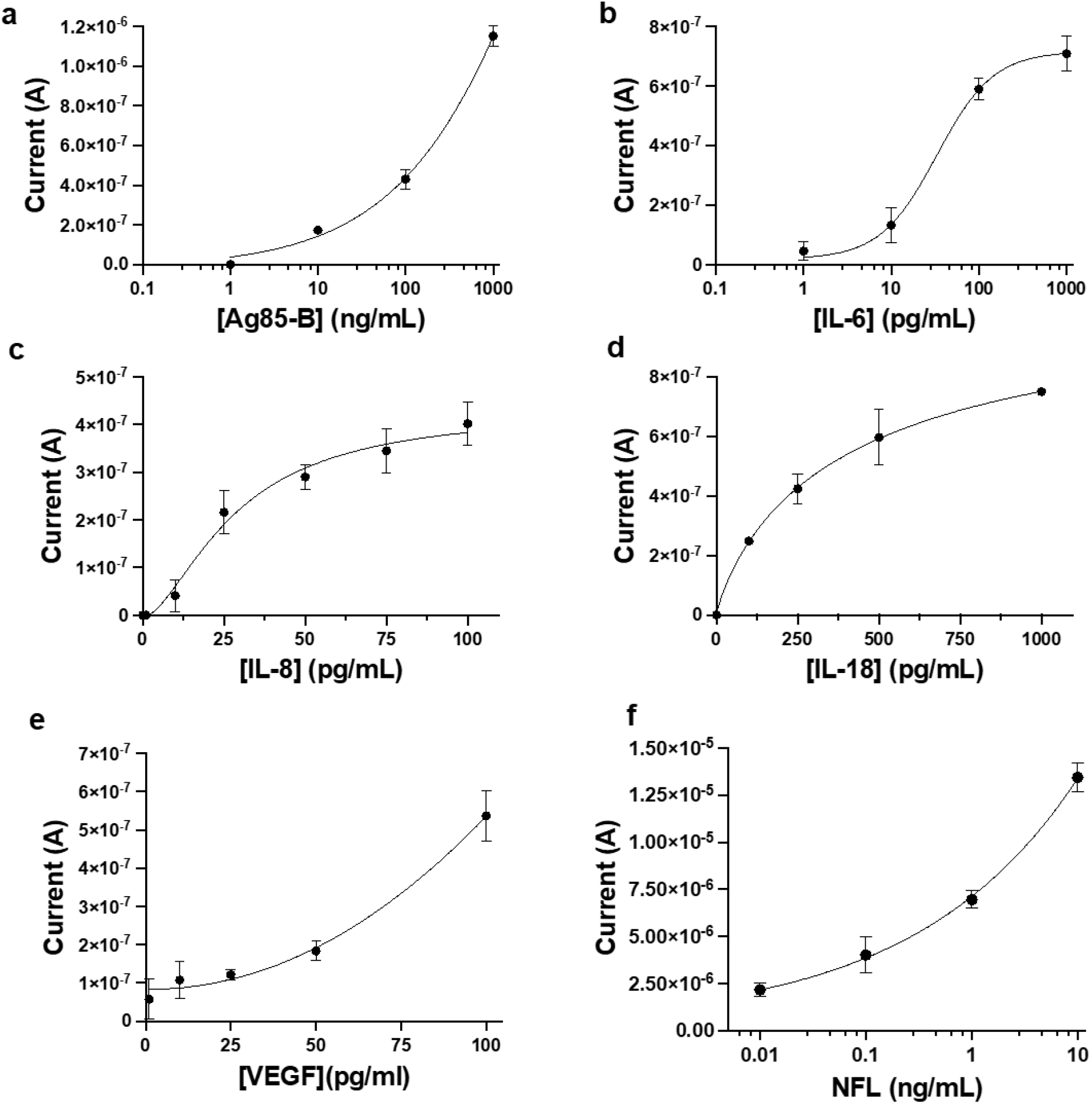
Calibration curves for different biomarkers using antifouling nanocomposite coated EC Biosensors. The left y-axis shows the current intensity for different concentrations of biomarkers run on EC biosensors using unprocessed human plasma. Different biomarkers tested include a) IL-6, b) IL-8, c) IL-8, d) VEGF, e) Ag85B, and f) PCT. Error bars represent the s.d. of the mean; n = 3. Analysis was done using 4-Parameter Logistic (4PL) curve fitting.

To explore the generality of this approach, we carried out a similar EC sandwich assay using specific capture antibodies for procalcitonin (PCT), a 116-amino acid peptide precursor for calcitonin which has a strong association with hepatitis C virus (HCV), sepsis, and other bacterial infections. We obtained LOD of 4 pg/mL, which is also more sensitive than the clinical cut-off value (**Table 1**, Supporting Information).^[10, 17]^ In addition, we performed a neurofilament light polypeptide (NF-L) assay using unprocessed whole blood on the Linxens sensor. NF-L is considered a promising biomarker for MS that has been shown to reflect disease activity in the clinical follow-up of the MS patients.^[18]^ With the Linxens sensor, a LOD of 0.3 pg/mL was obtained for NF-L using whole blood, which can be attributed to the highly efficient antifouling capability of our nanocomposite coating (**Figure S9**, Supporting Information).

### 2.3 Cross-reactivity of Antibody pair and Antigen

Cross-reactivity can be a significant issue in diagnostic immunoassays as it can result in over-or underestimation of sample analyte concentration.^[19]^ Thus, we performed a detailed cross-reactivity study for different pairs of MI and TBI antibodies and antigens, including NT-proBNP, in conventional plastic ELISA plates (**Figure 4a)**. Using an anti-NT-proBNP capture antibody with 10 ng/mL of the NT-proBNP analyte, we found that addition of multiple different secondary detection antibodies, including those directed against cTnI, BNP, S100, and glial fibrillary acidic protein (GFAP), failed to generate any detectable single compared to the blank, confirming that there is no non-specific binding of detection antibodies or cross-reactivity in this assay.

**Figure 4:**
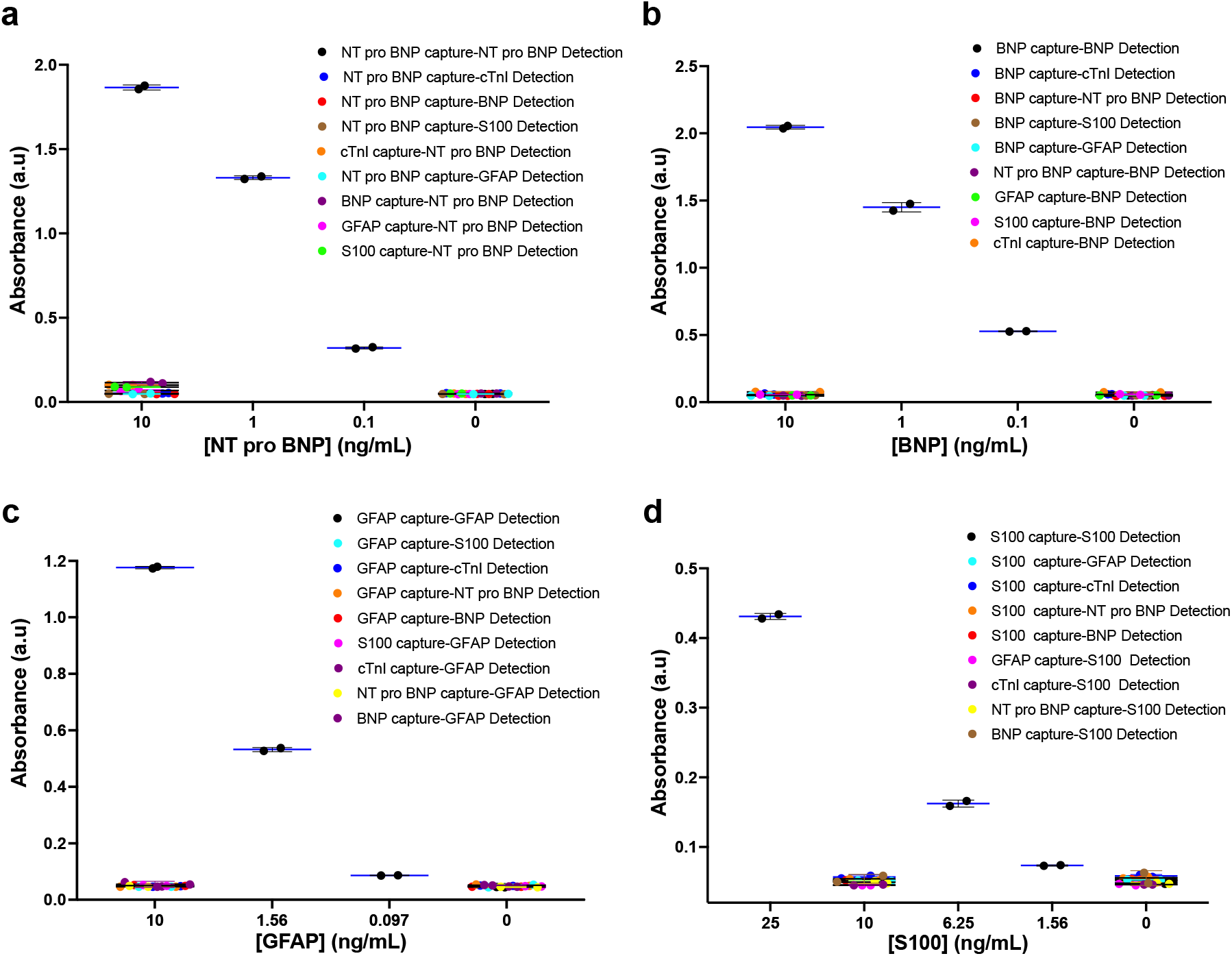
Specificity and cross-reactivity test for different biomarkers of Myocardial Infarction (MI) and Traumatic Brain Injury (TBI) done in 96 well plate. a) Specificity and cross-reactivity of NT-proBNP antigen against different non-specific capture and detection antibodies (anti-cTnITC, anti-BNP, anti-GFAP, and anti-S-100b) along with specific detection with anti-NT-proBNP capture and detection antibody at different concentrations of NT-proBNP. b) Specificity and cross-reactivity of BNP antigen against different non-specific capture and detection antibodies along with specific detection with anti-BNP capture and detection antibody at different concentrations of BNP. c) Specificity and cross-reactivity of GFAP antigen against different non-specific capture and detection antibodies along with specific detection with GFAP capture and detection antibody at different concentrations of GFAP. d) Specificity and cross-reactivity of S-100b antigen against different non-specific capture and detection antibodies along with specific detection with S-100b capture and detection antibody at different concentrations of GFAP.

We also explored whether the NT-proBNP analyte binds non-specifically to other capture antibodies (anti-cTnI, anti-BNP, anti-S100, and anti-GFAP) by coating ELISA plates with different capture antibodies followed by the addition of 10 ng/mL NT-proBNP and anti-NT-proBNP detection antibody. A minimum signal similar to blank was observed in each case (**Figure 4a**), again showing no cross-reactivity. Likewise, cross-reactivity of antibody pairs and antigens were also performed for BNP, GFAP, and S100 with all other capture and detection antibodies (**Figure 4b**,**c**,**d**). Concentration-based signals were observed only for specific antibody pairs, and no signal was observed using non-specific antibodies or target analytes; thus, these antibody pairs (NT-proBNP, BNP, GFAP, and s100) can be used for multiplexed detection in single assay.

For Troponin I, two antibody pairs (anti-cTnI and anti-cTnI-TC) were tested for cross-reactivity with other antibody pairs (BNP, NT-proBNP, GFAP, and s100) along with specific analytes, cTnI and cTnITC complex. We were able to demonstrate specific detection of cTnITC with an anti-cTnI antibody pair, which produced a lower specific signal and higher non-specific binding with BNP and a NT-proBNP capture antibody(**Figure S10a;** Supporting Information). We also obtained a concentration-dependent signal for cTnI with using anti-cTnI antibody pair, but observed very high non-specific binding of cTnI to BNP and NT-proBNP capture antibodies (**Figure S10b;** Supporting Information). Similar high specific signals were obtained for cTnITC using anti-cTnI-TC antibody pair, while there was almost no signal with the non-specific antibody pair (**Figure S10c**; Supporting Information). There was also very low specific binding of the anti-cTnI-TC antibody pair for cTnI (**Figure S10d**; Supporting Information). Thus, we can use the anti-cTnI-TC antibody pair (**Figure S10c**) with high specific signal for cTnITC and minimum cross-reactivity to other antibody pairs to create multiplexed assays for detection of MI and TBI biomarkers. In addition, we found that the assays for the cytokines IL-6 and IL-18 also only specifically react with their own capture and detection antibody pair and do not cross-react with antibody pairs against other interleukin types (IL-6, IL-8, and IL-18)(**Figure S11**; Supporting Information).

### 2.4 Multiplexed Detection using Plasma and Whole Blood

The high sensitivity and selectivity of the antifouling nanocomposite combined with the use of antibody pairs and corresponding target analytes that are free of any cross-reactivity made it possible to create a highly multiplexed EC sensor for simultaneous detection of multiple biomarkers both in plasma and whole blood. Different working electrodes in each EC sensor were individually functionalized with one of four different capture antibodies directed against cTnITC, S100, NT-proBNP, or anti-GFAP. Initially, we spiked plasma samples with different concentrations of cTnITC (0.01-10 ng/mL) followed by all four detection antibodies and observed a concentration-based signal only for cTnITC (**Figure 5a**), while minimal or no signals were observed for the other biomarkers (S100, NT-proBNP, and GFAP) or in unspiked (blank) plasma samples. We then spiked plasma samples with increasing concentrations of cTnITC (0.01-10 ng/mL) and decreasing concentrations of GFAP (10-0.01 ng/mL) and observed specific signal only for cTnITC and GFAP while no signal was observed for S100 and NT-proBNP along with blank samples (**Figure 5b**). We then carried out similar multiplexed EC sensor assays in human whole blood samples without any pre-treatment. In these studies, we were able to specifically detect cTnITC and GFAP (**Figure 5c)** as well as S100 and GFAP (**Figure 5d**), and develop calibration curves for each ligand simultaneously in multiplexed setting without observing any cross-reactivity with the capture or detection antibodies even in unprocessed whole blood.

**Figure 5:**
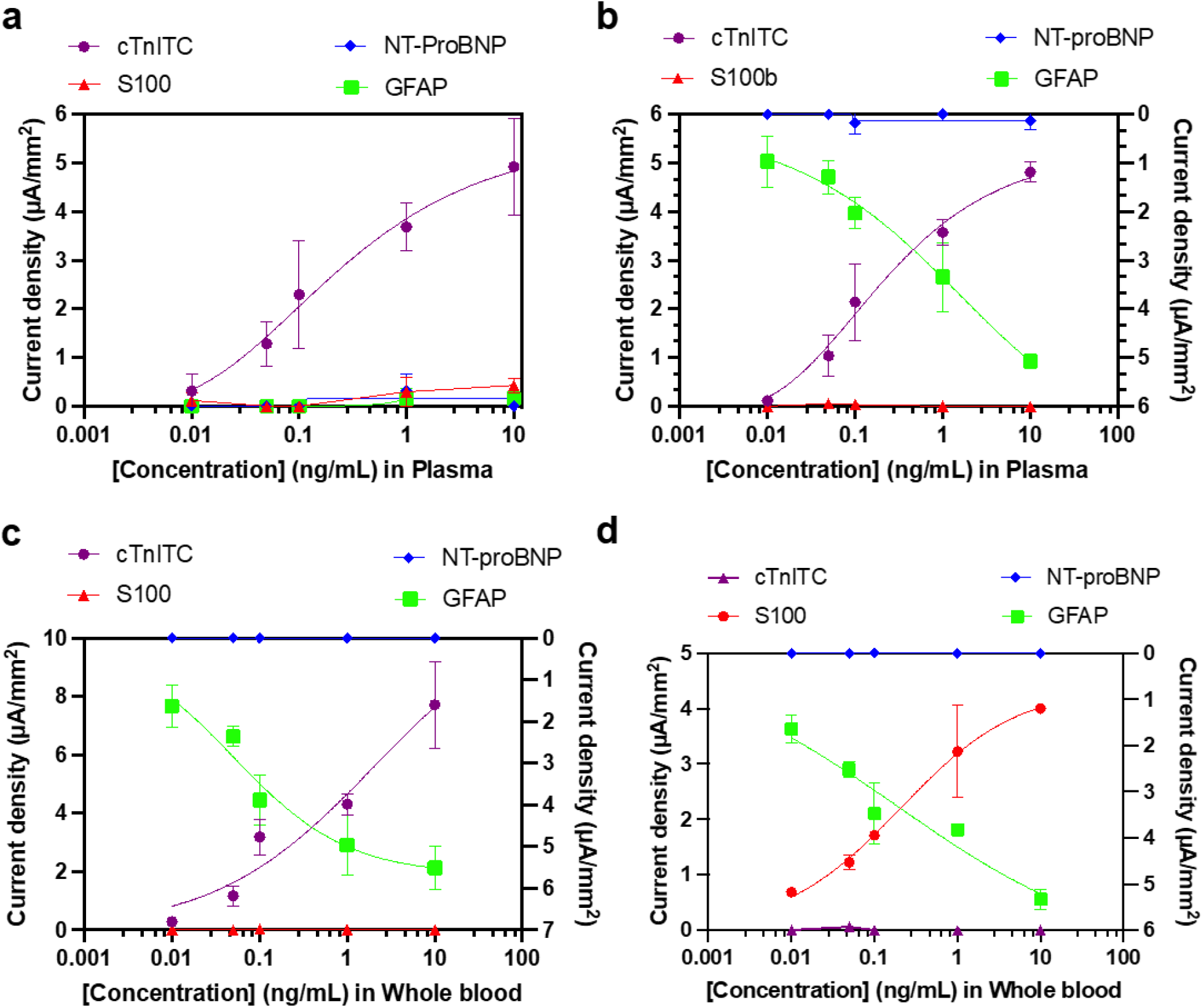
Specificity and Multiplexed detection for different biomarkers of MI and TBI using antifouling nanocomposite coated EC Biosensors. a) Calibration curve for multiplex detection of cTnITC using plasma sample on the EC Biosensor with four different capture antibodies on each electrode (anti-cTnITC, anti-S-100b, anti-GFAP, and anti-NT-proBNP). b) Calibration curve for multiplex detection of increasing concentration of cTnITC (left y-axis) and decreasing concentration of GFAP (right y-axis) using plasma sample (c) and whole blood (d) on the EC Biosensor with four different capture antibodies on each electrode. e) Calibration curve for multiplex detection of increasing concentration of s100 (left y-axis) and decreasing concentration of GFAP (right y-axis) using whole blood on the EC Biosensor with four different capture antibodies on each electrode. Error bars represent the s.d. of the mean; n = 3.

### 2.5 Detection of biomarkers in different complex biological fluids

For accurate disease diagnosis, a wide range of bodily fluids is employed in clinical settings, such as blood, serum, saliva, sputum, and sweat. However, the inherent biological and physiochemical properties of these complex fluids often hinders clinical diagnosis. The most common problem is that molecules present within these fluids can interact with the analyte or with the sensor surface, which leads to the reduction of the signal.^[20]^ The sensitivity of EC sensors also can be reduced due to limited access of redox molecules or analytes to the sensor surface in these samples that can be highly viscous and rich in cells as well as molecules. Futhermore, most clinical biomarkers are present at very low concentrations (fM to pM range) in these samples, and they can often cross-react with the large molecules, such as albumin and immunoglobulins, which are present at much higher concentration (μM to mM range).^[21]^ Biological samples are also unstable, and often environmental triggers have an impact in antigen–antibody interaction, non-specific binding, and degradation of sample.^[21]^ Thus, the detection of multiple biomarkers in clinical samples can provide different challenges due to the complexities of the matrix.^[3]^ Given these observations, as the LOD is one of the critical parameters in clinical bioassay, it must be measured in the complex fluid sample intended for use using the complete sample preparation sequence required to carry out the assay.^[20-22]^

We have tackled these issues by developing a highly efficient antifouling coating, carrying out rigorous screening of antibodies, and optimizing assay development. To validate the nanocomposite’s antifouling effect and show that our EC biosensor can perform the assay in different complex biological fluids, we compared the efficiency of a single-step sandwich assay for GFAP in buffer, plasma, and whole blood (**Figure 6a**). As expected, we observed a similar calibration curve in the range of 0.01 to 10 ng/mL in all three samples (LOD of GFAP in buffer, plasma, and whole blood was 16, 7, and 2 pg/mL, respectively). The fact that the EC biosensor with antifouling coating works similarly in unprocessed whole blood as well as plasma was also demonstrated by showing that the calibration curve for NF-L in whole blood and plasma closely overlap each other, with LODs of 10 and 3 pg/mL, respectively (**Figure 6b)**.

**Figure 6:**
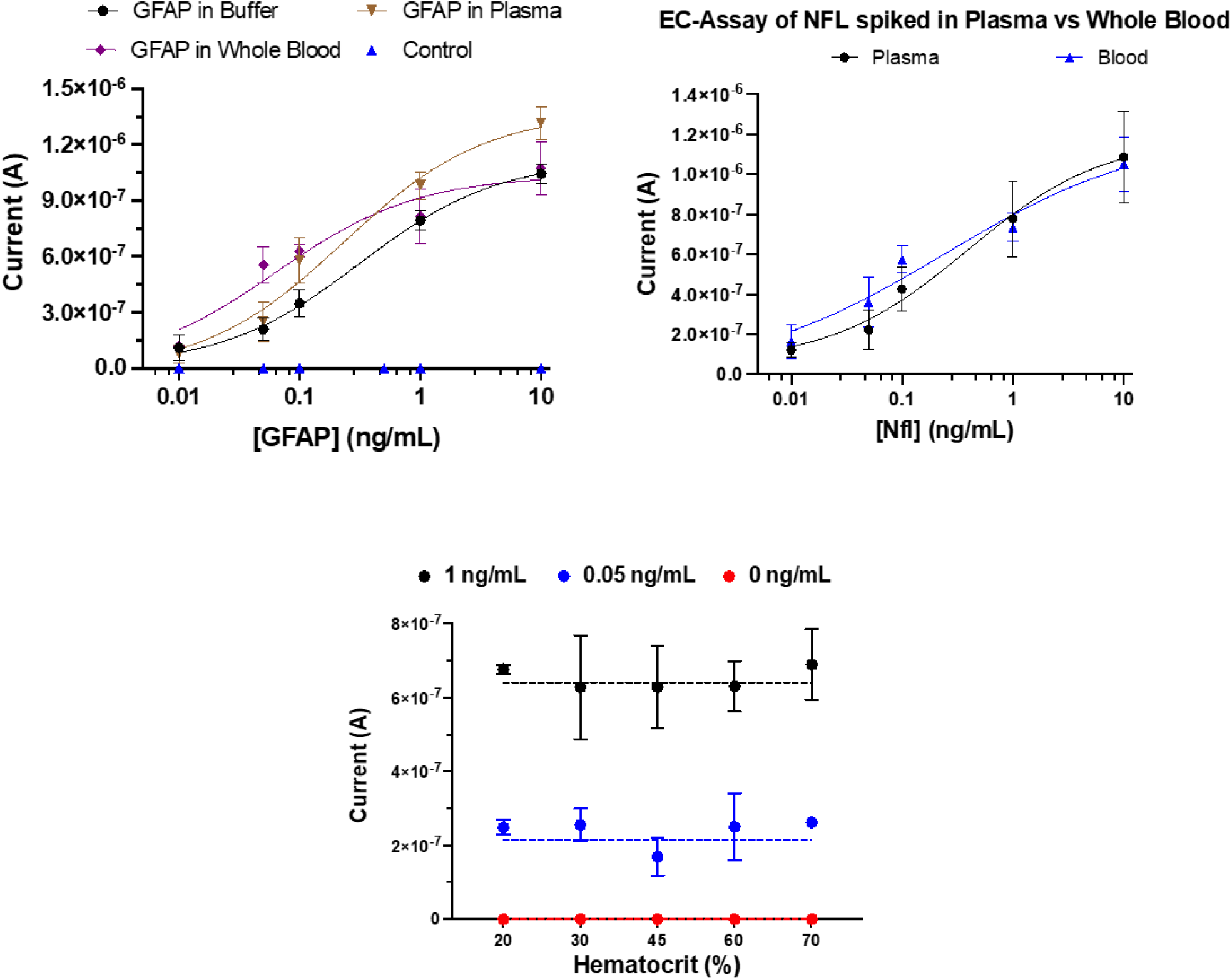
Matrix effect for detection of different biomarkers on EC Biosensors. a) Calibration curve of GFAP using buffer (black dot), plasma (orange), and whole blood (purple) on EC Biosensor. b) Calibration curve of NF-L using plasma (black dot) and whole blood (blue) on EC Biosensor. c) Detection of different concentration of NF-L (1, 0.05, and 0 ng/mL) at different % of hematocrit (20%-70%). Line graph shows the mean current density for the assay of NF-L using plasma sample on EC biosensor. Error bars represent the s.d. of the mean; n = 3.

### 2.6 Effect of hematocrit levels

Hematocrit levels (erythrocyte concentrations) in blood fluctuate between individuals and various disease states, and they can impact the accuracy of biomarker detection in whole blood samples.^[23]^ We therefore measured the effect of different hematocrit levels (20%, 30%, 45%, 60%, and 70%) on the performance of the EC assay using a plasma sample as a control. When we spiked these samples with two different concentrations of NF-L (0, 0.1, and 1 ng/mL) and blank, there were no significant difference in the signals observed between samples with different hematocrits or in the plasma sample (**Figure 6c**).

## 3. Discussion

In this study, we described development methods and optimization strategies for creation of multiplexed EC sensors that can be used to detect clinically relevant biomarkers in complex biological fluids, such as unprocessed whole blood and plasma, with high sensitivity and specificity as well as minimal cross reactivity. The optimization of individual components in sandwich immunoassays combined with the high efficiency antifouling coating enabled highly sensitive detection with near-zero cross-reaction for selective biomarkers for MI, TBI, MS, and TB when present in the clinically relevant range within plasma and blood samples. The ability of these sensors to simultaneously detect multiple different biomarkers should allow for more accurate detection and evaluation of diseases with a limited quantity of clinical samples, in addition to offering the possibility of early diagnosis at POC.

We achieved multiplexing of four different biomarkers in the same sensor by antibody screening, assay parameter optimization, and carrying out cross-reactivity studies. Potential cross-reactivity was tested by replacing the actual antigen and antibodies (capture and detection) with similar size proteins and non-specific antibodies. We consistently observed specific signals for the real analytes with minimum readout when using non-specific molecules, which allowed us to delineate the selectivity and also demonstrate superiority in terms of the accuracy of detection. For instance, using NT-proBNP as cardiovascular disease protein biomarker, cross-reactivity with non-specific protein biomarkers (BNP, cTnI, S100, and GFAP) was assessed using similar sized antibodies and complex protein structures. When the capture antibodies targeting the analytes were replaced with antibodies against other targets, the output signals were consistently low or zero. In addition, the use of locally precipitating TMB allowed us to build versatile multiplexed biosensors in close juxtaposition, each using a different antigen-antibody pair, which do not cross-react.

EC sensors are not used often for clinical diagnosis or POC application largely because they are prone to electrode fouling when used with complex biological fluids. By integrating an antifouling nanocomposite coating, we were able to detect multiple disease biomarkers in complex samples, such as human whole blood and plasma, and still obtain clinically relevant LODs. Our studies on detection of the neurological disease biomarkers, GFAP and NF-L, also demonstrated that hematocrit levels in blood had no significant effect on our EC biosensor with antifouling coating. We expect that other biomarkers will show similar behaviors, but a detailed study should be carried out with each biomarker explored in the future. Moreover, the same antifouling coating worked equally well when integrating different kinds of EC sensors, including screen printed electrodes and transducer-like materials. Thus, this antifouling coating is a key feature of our EC sensors that provides a significant advantage that is highly relevant for clinical diagnostics as well as POC applications.

Finally, as a proof-of-concept, we leveraged our multiplexed EC sensor to develop a TB diagnostic. Nearly 10 million people develop TB every year and this results in approximately 1.5 million deaths worldwide each year even though it is entirely curable with early diagnosis and an effective treatment plan.^[24]^ Current TB diagnostic tests are either difficult to access as they are only available in hospital settings or they have low accuracy, reliability, yield delayed results, and require expertise and specialized facilities. In addition, sputum sample collection from suspected TB patients is cumbersome, exacerbating the difficulty in making a timely diagnosis. An accurate POC device that could diagnose TB early and guide the use of medical treatment would significantly improve the management of this disease in endemic areas, decreasing morbidity and slowing disease transmission.^[25]^ A combination of TB disease-specific biomarker Antigen (Ag)85B (Ag85-B) and host response cytokines IL-6, IL-8, IL-18, and VEGF were previously combined to configure a blood-based triage test for active pulmonary TB based on analysis of an extensive cohort study.^[15]^ Importantly, the performance of this biomarker panel was validated by an independent blinded set of samples, and it meets the World Health Organization’s (WHO) mimimal target product profile (TPP) for a blood-based TB triage test.

Thus, we believe that this new approach for fabricating multiplexed EC sensors can be used to develop POC diagnostics that could have near-term impact on healthcare world-wide. In summary, in this study, we have addressed four significant challenges including, i) prevention of biofouling from the complex biological samples like blood, ii) choosing a coherent group of biomarkers that is highly specific to a disease condition, iii) developing sensitive assay without any cross-reaction, and iv) integrating the technology into a multiplexed EC sensor. Combined, this approach enable rapid and cost-effective development of multiplexed biosensors that provide highly sensitive and specific detection of various clinically relevant biomarkers for complex diseases, as we demonstrated for TB, MI, and TBI. They also can be adapted to detect any desired analyte if appropriate specific capture and detection molecules (e.g., antibodies, aptamers, etc.) are available. Thus, these multiplexed EC sensor arrays could provide a new approach to disease diagnosis as well as environmental monitoring in POC settings.

## 4. Methods

### 4.1 Fabrication of Electrochemical (EC) sensor

The EC sensor with gold electrodes which was purchased from Telic Company was custom fabricated using a standard photolithography process. The chips were cleaned by sonicating in acetone followed by isopropyl alcohol and cleaned with plasma cleaner for 8 min as described previously.^[8b]^ Antifouling coating solution consisting of prGOx and BSA crosslinked with GA was drop-casted over the chips at 85 °C for 45s.^[8a]^ The chips were then washed by dipping in PBS immediately at 400 rpm for 10 min.

### 4.2 Conjugation of Capture Antibodies

After drying the coated chips with a slide spinner (Millipore Sigma, no. 674 664), 400 mM EDC (Thermo Fisher Scientific, no. 22 980) and 200 mM NHS (Sigma-Aldrich, no. 130 672) were dissolved in 0.05 M MES (2-(N-morpholino)ethanesulfonic acid) Buffer (pH 6.2) and deposited over the chips for 30 min at room temperature in dark. The chips were then quickly rinsed with Milli Q water and dried with compressed air followed by spotting of optimized concentration of capture antibody on top of three working electrodes and BSA (5 mg/mL) over the 4^th^ electrode as a negative control using Xtend capillary microarray Pin (LabNEXT, no. 007-350). The chips spotted with capture antibody and BSA were stored overnight at 4 °C in a humidity chamber followed by washing with PBS. The chips were then exposed to 1 M ethanolamine (Sigma-Aldrich, USA, no. E9508) in PBS to quench the unreacted glutaraldehyde groups for 30 min and blocked with 10 μL of 2.5% BSA in PBS for 1 h.

### 4.3 Detection of Biomarkers in EC Sensor

Detection of different biomarkers on the EC biosensor was performed using the optimized conditions. Three working electrodes were spotted with respective capture antibodies diluted in PBS [anti-BNP (HyTest, no. 50E1cc), anti-NT-proBNP (Medix Biochemica, no. 100 521), anti-cTnI (Abcam, no. ab243982), anti-cTnI-TC (Advanced ImmunoChemical, no. 2-TIC-rc) anti-GFAP (HyTest, no. GFAP83cc), anti-S100b (HyTest, no. 8B10cc)], anti NF-L (Uman Diagnostics, no. 27016), anti-IL-6 (Abcam, ab246838), anti-IL-8 (Abcam, ab215402), anti-IL-18 (Abcam, ab218185), anti-VEGF (R&D system DY493-05), and anti-Ag85-B (Abcam, ab36731). Antigens were then spiked into the plasma samples at different concentrations ranging from 1 pg/mL to 10,000 pg/mL and mixed with the optimized concentration of biotin conjugated detection antibody in the ratio of 9:1. Different antigens used includes [BNP-32 (Bachem, no. 4 095 916), NT-proBNP (Medix Biochemica, no. 610 090), cTnI (Medix Biochemica, no. 610 102), cTnI-TC complex (HyTest, no. 8T62), GFAP (HyTest, no. 8G45), S100b (HyTest, no. 8S9h), NF-L (Encor, no. PROT-r-NF-L), IL-6, IL-8, IL-18, VEGF, and Ag85-B, respectively. 15 μL of the sample detection antibody mixture was then added to the EC biosensor and incubated with agitation at 400 rpm for 30 min (single-step assay). For 2 step assay, 15 μL of the sample antigen was incubated for 1 h followed by washing and the addition of 10 μL of detection antibody for 15 min. Conjugation of biotin to detection antibody was done using Biotin Conjugation Kit (Fast, Type A)– Lightning-Link (Abcam, USA, no. ab201795) using manufacturer’s protocol except for anti-NF-L detection antibody which was already linked with biotin when purchased. Detection antibodies used for the assay include anti-BNP (HyTest, no. 24C5cc), anti-NT-proBNP (Medix Biochemica, no. 100 712), anti-cTnI (Abcam, no. ab243982), anticTnI-TC (Advanced ImmunoChemical, USA no. 2-TC), anti-GFAP (HyTest, no. GFAP81cc), anti-S-100b (HyTest, no. 6G1cc), anti NF-L (Uman Diagnostics, no. 27018), anti-IL-6, anti-IL-8, anti-IL-18, anti-VEGF, and anti-Ag85-B, respectively. After 30 min incubation, the EC biosensors were washed with PBST (PBS with 0.05% Tween 20 (Sigma-Aldrich, no. P9416)). 2-5 μg/mL of spHRP (Thermo Fisher Scientific, no. N200) diluted in 0.1% BSA in PBST was then added to each EC biosensor for 5 min followed by washing and addition of precipitating TMB, Sigma-Aldrich, USA, no. T9455) membrane substrate for 2 min. The EC biosensors were finally washed with PBST before taking the electrochemical measurements in PBST using a potentiostat by a CV with a scan rate of 1 V/s between −0.5 and 0.5 V versus on-chip integrated gold quasi reference electrode. Linxens sensor was cleaned by dipping in 50 mg/mL sodium carbonate for 10s followed by rinsing in water and dipping in 920 mM sulfuric acid for another 10s before final rinse with water. Gii-sens electrode were directly used without cleaning. Linxens and Gii-sens electrode were characterized by measuring CV in PBS containing 5 mM [Fe(CN)_6_]^3-/4-^ at 200 mV/s between -0.5 to 0.5V. All samples were collected under the approval of the Institutional Review Board for Harvard Human Research Protection Program (IRB21-0024).

### 4.4 Cross-reactivity

Specificity of Antigen and Antibody was performed in Nunc™ MaxiSorp™ ELISA plates (BioLegend, no. 423501). For each biomarker four different concentrations were run with specific antibody pairs to observe the signal for specific binding. To see if there is any non-specific binding of antigen to capture antibody of other biomarkers, all the non-specific capture antibodies were coated to the plates followed by the addition of high concentration analyte (10 ng/mL) and negative control (0 ng/ml) and detection antibody for the analyte. To test non-specific binding between the antigen and detection antibody of other biomarkers, a specific capture antibody was coated to the plate and a high concentration analyte (10 ng/mL) and negative control were added to the plate followed by all other non-specific detection antibodies. All the assays were performed in buffer (1 % BSA in PBS). For instance, in case of BNP specificity test, four concentrations of BNP (0, 0.1, 1, and 10 ng/mL) (black dots, **Figure 4b**) were tested with specific antibody pair for BNP. For non-specific capture antibody-antigen binding test, four different capture antibodies (anti-NT-pro BNP, anti-cTnI, anti-GFAP, and anti-S100) were coated at 1 μg/mL followed by the addition of either 10 ng/mL or 0 ng/mL of BNP. After washing anti-BNP detection antibody was added followed by Streptavidin-HRP and TMB (ThermoScientific, no. 34022). Similarly, for the non-specific antigen-detection antibody binding test, BNP capture antibody followed by 0 or 10 ng/mL of BNP was added. After washing, non-specific biotinylated detection antibodies (anti-NT-proBNP, anti-cTnI, anti-GFAP, and anti-S100) were added followed by Streptavidin-HRP and TMB. Likewise, specificity test for other biomarkers including NT-proBNP, GFAP, and S100 was done in similar way with specific and non-specific antigen-antibody pairs. Similarly, for cTnI and cTnITC specificity test was performed with both Abcam antibody pair (specific to cTnI) and Advanced ImmunoChemical antibody pair (specific to cTnITC). Likewise, cross-reactivity test was also performed for different biomarkers of TB including Il-6, IL-8, and IL-18.

### 4.5 Multiplexed detection of Biomarkers in EC Platform

For the detection of cTnITC in multiplexed EC biosensor, four different capture antibodies (anti-cTnITC, anti-S100, anti-NT-proBNP, and anti-GFAP) were spotted on four different electrodes of the chip at 500 μg/mL. Increasing concentration of cTnITC (0, 0.01, 0.05, 0.1, 1, and 10 ng/mL) spiked in plasma samples were mixed with optimum concentration of all four biotinylated detection antibodies and added to the EC biosensor for 30 min. Chips were washed and spHRP was added for 5 min followed by TMB for 2min before washing and reading the chips. In the next experiment for parallel detection of cTnITC and GFAP, all capture antibodies were spotted as earlier. Increasing concentration of cTnITC and decreasing concentration of GFAP (0.01 cTnITC + 10 GFAP; 0.05 cTnITC +1 GFAP; 0.1 cTnITC + 0.1 GFAP; 1 cTnITC + 0.05 GFAP; 10 cTnITC + 0.01 GFAP) spiked in plasma samples was mixed with all four biotinylated detection antibody and incubated for 30 min. Likewise, simultaneous multiplexed detection of cTnITC and GFAP was performed in unprocessed whole blood with increasing concentration of cTnITC and decreasing concentration of GFAP (0.01 cTnITC + 10 GFAP; 0.05 cTnITC +1 GFAP; 0.1 cTnITC + 0.1 GFAP; 1 cTnITC + 0.05 GFAP; 10 cTnITC + 0.01 GFAP). Similarly, simultaneous multiplexed detection of S100 and GFAP in the whole blood sample was also performed.

### 4.6 Matrix and hematocrit effects

The effect of different types of matrices on the performance of EC biosensor was evaluated by running the calibration curve of GFAP using buffer (2.5 % BSA in PBS), plasma, and whole blood sample with the optimized conditions. The antifouling effect of different matrix components was further evaluated by running a calibration curve of NF-L in plasma vs unprocessed whole blood. To study the hematocrit effect, whole blood samples collected in Sodium heparin tube was used to separate plasma from RBCs. Briefly, Heparinized tubes were centrifuged for 10 minutes at 1,500 x g using a refrigerated centrifuge. Following centrifugation, plasma was immediately transferred into a clean polypropylene tube using a pipette and kept over ice. Blood samples with different nominal hematocrit values in the range of 20 to 70% were then prepared by mixing different ratios of plasma and RBCs followed by gentle mixing. EC assay of NF-L was immediately performed after the preparation of plasma and whole blood with different hematocrit levels.

### 4.7 Statistical Analysis

ELISA reading is reported as absorbance (a.u.) of the mean of replicates and error bars represent the standard deviation (s.d.) of the mean; n = 2. For EC biosensor studies, peak heights were calculated using Nova 1.11 software. For data analysis peak height (μA) was converted to current density (μA/mm^2^) for further analysis by dividing the peak height by the surface area of the working electrode (0.1576 mm^2^). Error bars represent mean ± s.d. for all EC biosensor studies (sample sizes and statistical tests used are indicated in the Figure legends). All data were plotted, and statistical tests were performed using GraphPad Prism 8, and 4-Parameter Logistic (4PL) curve fitting was done for calibration curve analysis.

## Data Availability

All data produced in the present study are available upon reasonable request to the authors

## Supporting Information

Supporting Information is available from the Wiley Online Library or from the author. Supporting Information includes optimization of coating time, TMB stability, Linxens and Gii-sens characterization, two-step calibration curves, optimization studies, cross-reactivity studies, and calibration curve.

## Acknowledgments

We would like to acknowledge Linxens and Integrated Graphene for providing the electrodes for the study. This work was supported by the Wyss Institute for Biologically Inspired Engineering at Harvard University. MR was supported from Internal funding from bioengineering at UTD

## Author contributions

SS, MR, ND conceived the study under the guidance of RA, PJ, and DEI. MR under the guidance of RA and PJ contributed to TB-related work. Experiments were performed and validated by SS, MR, and ND. All authors contributed to discussion, manuscript preparation, and editing.

## TOC figure

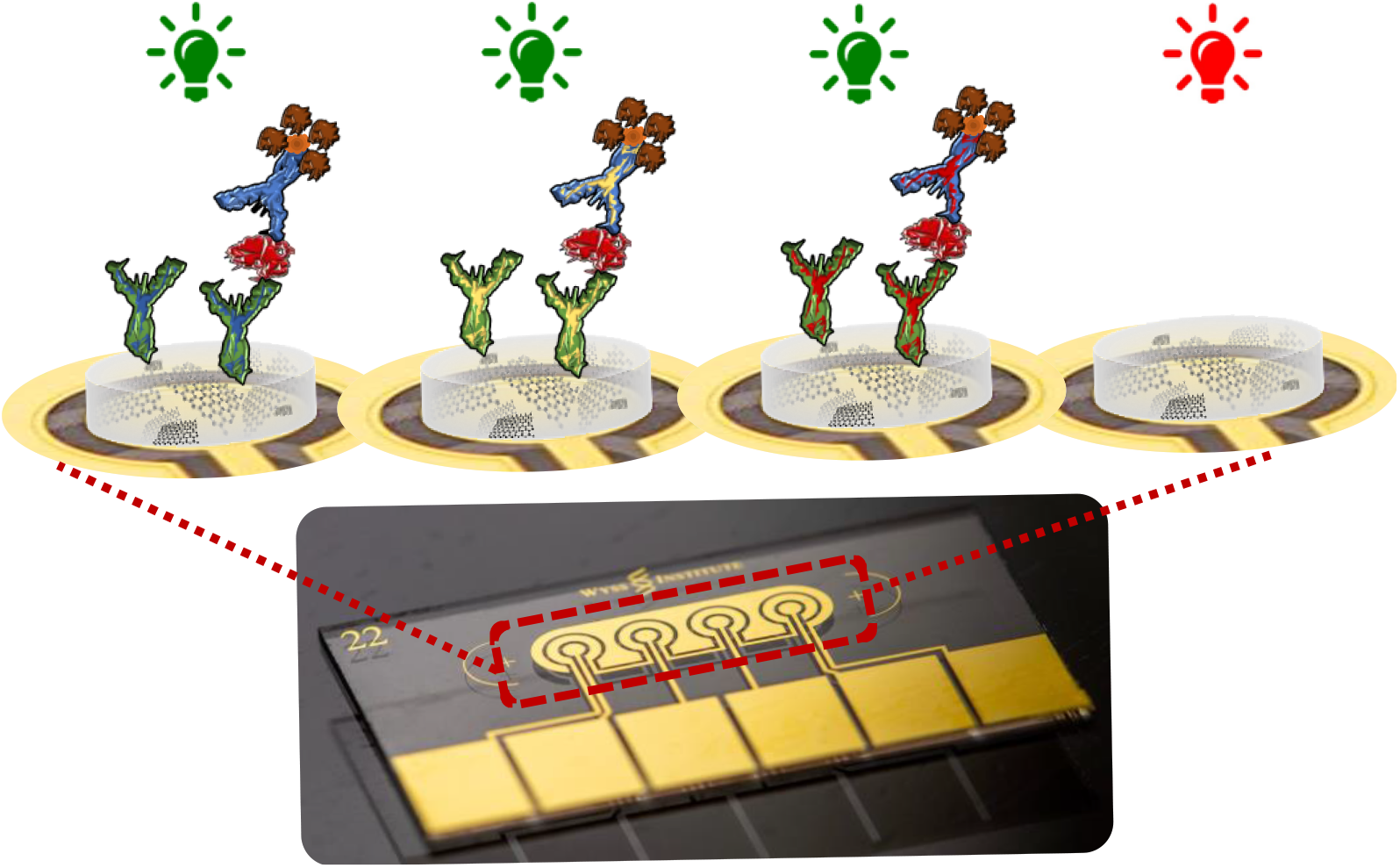

## REFERENCES

[1] J. Yang, K. Wang, H. Xu, W. Yan, Q. Jin, D. Cui, Talanta 2019, 202, 96.

[2] X. Li, Y. Zhang, B. Xue, X. Kong, X. Liu, L. Tu, Y. Chang, H. Zhang, Biosensors and Bioelectronics 2017, 92, 517.

[3] Y. Rosenberg-Hasson, L. Hansmann, M. Liedtke, I. Herschmann, H. T. Maecker, Immunologic research 2014, 58, 224.

[4]a) A. Kaushik M. A. Mujawar, Vol. 18, Multidisciplinary Digital Publishing Institute, 2018, 4303;

b) K. S. Prasad, X. Cao, N. Gao, Q. Jin, S. T. Sanjay, G. Henao-Pabon, X. Li, Sensors and Actuators B: Chemical 2020, 305, 127516;

c) C. Dincer, R. Bruch, A. Kling, P. S. Dittrich, G. A. Urban, Trends in biotechnology 2017, 35, 728.

[5] S. S. Timilsina, P. Jolly, N. Durr, M. Yafia, D. E. Ingber, Accounts of Chemical Research 2021, 54, 3529.

[6]a) N. Arroyo-Currás, P. Dauphin-Ducharme, K. Scida, J.L. Chávez, Analytical Methods 2020, 12, 1288;

b) S. Campuzano, M. Pedrero, M. Gamella, V. Serafín, P. Yáñez-Sedeño, J.M. Pingarrón, Sensors 2020, 20, 3376.

[7] U. Zupančič, P. Jolly, P. Estrela, D. Moschou, D. E. Ingber, Advanced Functional Materials 2021, 31, 2010638.

[8]a) S. S. Timilsina, N. Durr, M. Yafia, H. Sallum, P. Jolly, D. E. Ingber, Advanced healthcare materials 2021, e2102244;

b) J. Sabaté del Río, O. Y. Henry, P. Jolly, D. E. Ingber, Nature nanotechnology 2019, 14, 1143.

[9]a) S. T. Sanjay, M. Dou, J. Sun, X. Li, Scientific reports 2016, 6, 1;

b) M. Dou, S. T. Sanjay, D. C. Dominguez, S. Zhan, X. Li, Chemical communications 2017, 53, 10886;

c) X. Wei, W. Zhou, S. T. Sanjay, J. Zhang, Q. Jin, F. Xu, D. C. Dominguez, X. Li, Analytical chemistry 2018, 90, 9888.

[10] I. Taneja, G. L. Damhorst, C. Lopez‐Espina, S. D. Zhao, R. Zhu, S. Khan, K. White, J. Kumar, A. Vincent, L. Yeh, Clinical and translational science 2021, 14, 1578.

[11] M. M. Ling, C. Ricks, P. Lea, Expert review of molecular diagnostics 2007, 7, 87.

[12] N. Madaboosi, R. R. Soares, V. Chu, J. P. Conde, Analyst 2015, 140, 4423.

[13]a) M. M. Redfield, R. J. Rodeheffer, S. J. Jacobsen, D. W. Mahoney, K. R. Bailey, J. C. Burnett, Journal of the American College of Cardiology 2002, 40, 976;

b) T. Mueller, M. Egger, E. Peer, E. Jani, B. Dieplinger, Clinica Chimica Acta 2018, 487, 66.

[14] D. C. Malaspina, G. Longo, I. Szleifer, PLoS One 2017, 12, e0185518.

[15] R. Ahmad, L. Xie, M. Pyle, M. F. Suarez, T. Broger, D. Steinberg, S. M. Ame, M. G. Lucero, M. J. Szucs, M. MacMullan, Science translational medicine 2019, 11, eaaw8287.

[16] C. T. Turner, R. K. Gupta, E. Tsaliki, J. K. Roe, P. Mondal, G. R. Nyawo, Z. Palmer, R. F. Miller, B. W. Reeve, G. Theron, The Lancet Respiratory Medicine 2020, 8, 407.

[17]a) J. Dinulos, Habif’s Clinical Dermatology. 7th ed. Philadelphia, PA: Elsevier 2021;

b) N. I. Rashwan, M. H. Hassan, Z. M. M. El-Deen, A. El-Abd Ahmed, Pediatrics & Neonatology 2019, 60, 149.

[18] K. N. Varhaug Ø Torkildsen, K.-M. Myhr, C. A. Vedeler, Frontiers in neurology 2019, 10, 338;

b) N. Siller, J. Kuhle, M. Muthuraman, C. Barro, T. Uphaus, S. Groppa, L. Kappos, F. Zipp, S. Bittner, Multiple Sclerosis Journal 2019, 25, 678.

[19] J. Tate, G. Ward, The clinical biochemist reviews 2004, 25, 105.

[20] M. L. Chiu, W. Lawi, S. T. Snyder, P. K. Wong, J. C. Liao, V. Gau, JALA: Journal of the Association for Laboratory Automation 2010, 15, 233.

[21] M. A. Johansson, K.-E. Hellenäs, Analyst 2004, 129, 438.

[22] J. Sun, Y. Liu, Micromachines 2018, 9, 142.

[23]a) P. Denniff, N. Spooner, Bioanalysis 2010, 2, 1385;

b) G. N. Eick, P. Kowal, T. Barrett, E. A. Thiele, J. J. Snodgrass, Biodemography and Social Biology 2017, 63, 116.

[24] A. MacNeil, P. Glaziou, C. Sismanidis, A. Date, S. Maloney, K. Floyd, Morbidity and Mortality Weekly Report 2020, 69, 281.

[25] K. Dheda, M. Ruhwald, G. Theron, J. Peter, W. C. Yam, Respirology 2013, 18, 217.

