## Supplementary Information for "Biofabrication of multiplexed electrochemical immunosensors for simultaneous detection of clinical biomarkers in complex fluids"

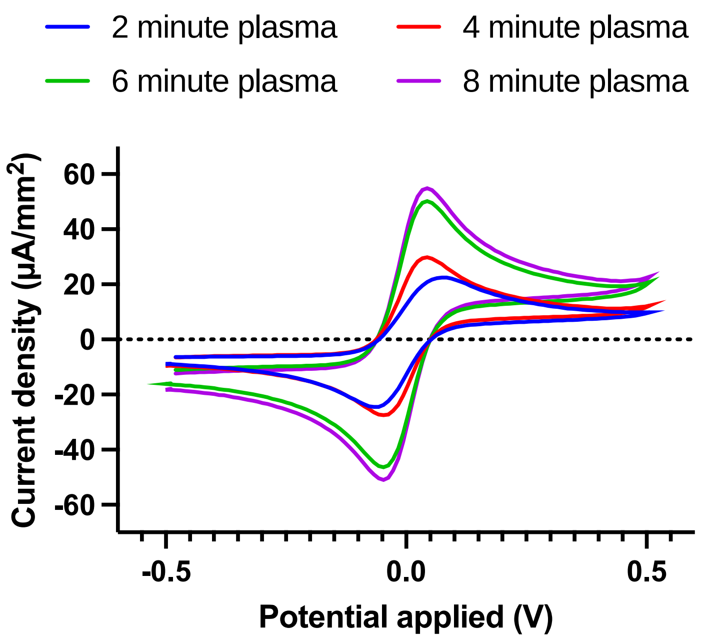


**Figure S1:** Optimization of plasma cleaning time for Sensor before application of coating. Typical voltammograms showing oxidation and reduction peak of ferri-/ferrocyanide with different plasma treatment time.

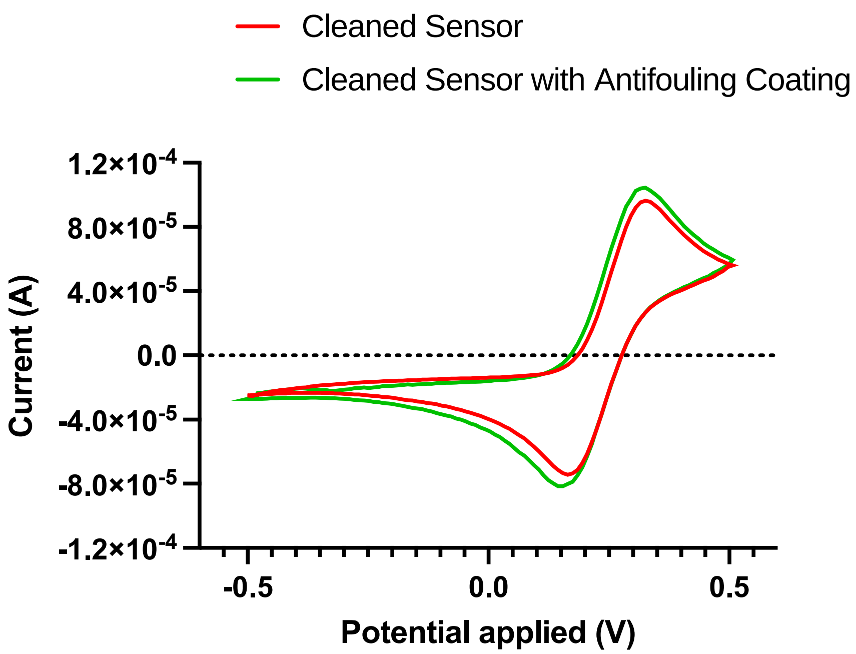


**Figure S2:** Characterization of Linxens sensor. Cyclic voltammetry showing oxidation and reduction peak of ferri-/ferrocyanide of Linxens sensor before and after coating the sensor with anti-fouling nanocomposite.

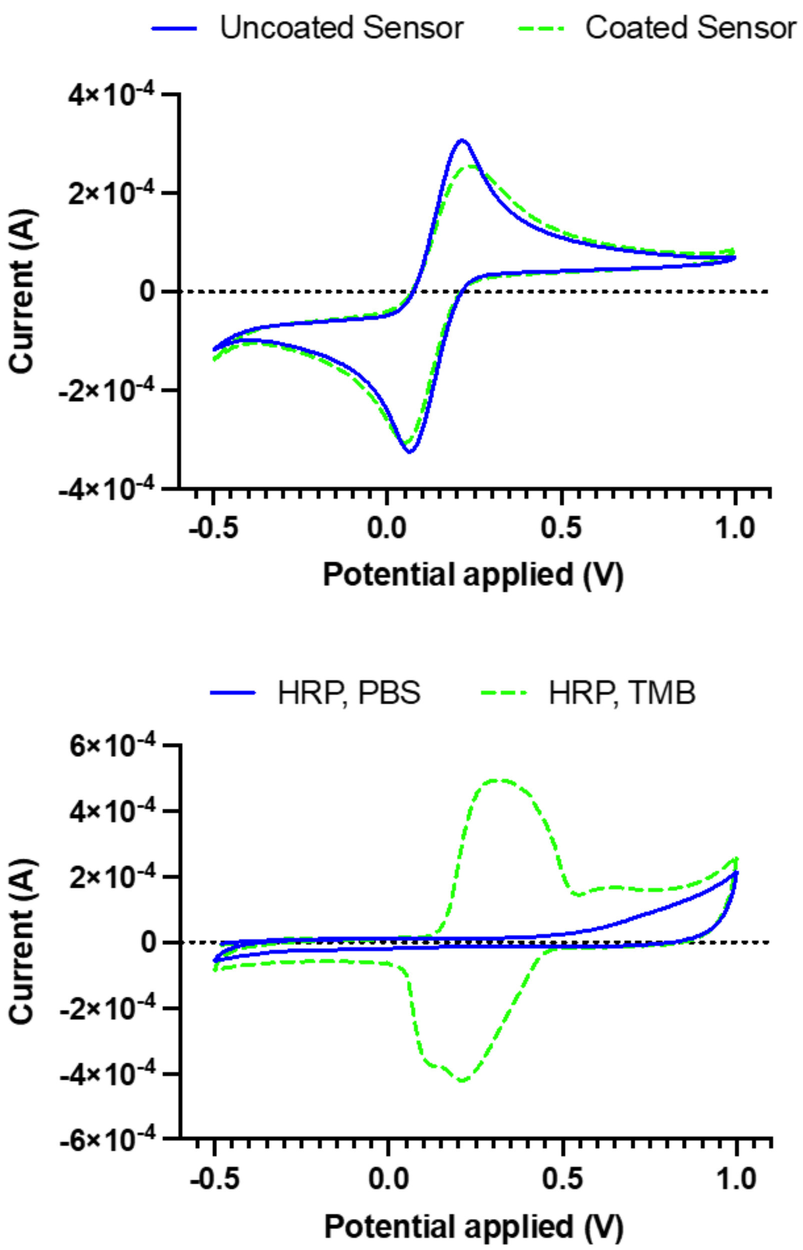


**Figure S3:** Characterization of Gii-sens sensor. a) Cyclic voltammetry of Gii-sens showing oxidation and reduction peak of ferri-/ferrocyanide sensor before and after coating the sensor with anti-fouling nanocomposite. b) Cyclic voltammetry showing oxidation and reduction peak of oxidixed TMB of HRP modified Gii-sens sensor after the addition of TMB (positive control) and PBS (negative control).

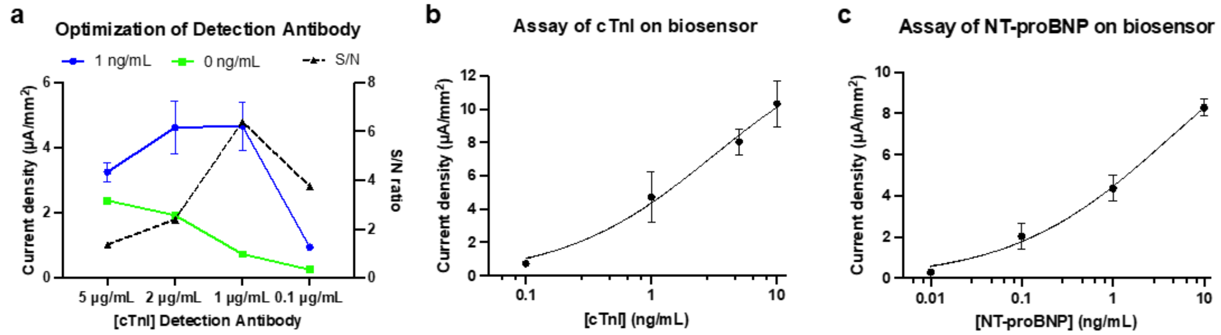


**Figure S4:** Optimization and two step assay for different biomarkers. a) Optimization of detection antibody for the assay of cTnI. Left y-axis show current density for 1 ng/mL (blue) and 0 ng/mL (green) of cTnITC while right y-axis shows signal to noise ratio at different concentration of anti-cTnI detection antibody. b) Calibration curve of cTnI run on the EC Biosensor with antifouling coating using two step assay and optimized detection antibody concentration. c) Calibration curve of NT-proBNP run on the EC Biosensor with antifouling coating using two step assay and similar assay conditions. Error bars represent the s.d. of the mean, n=3.

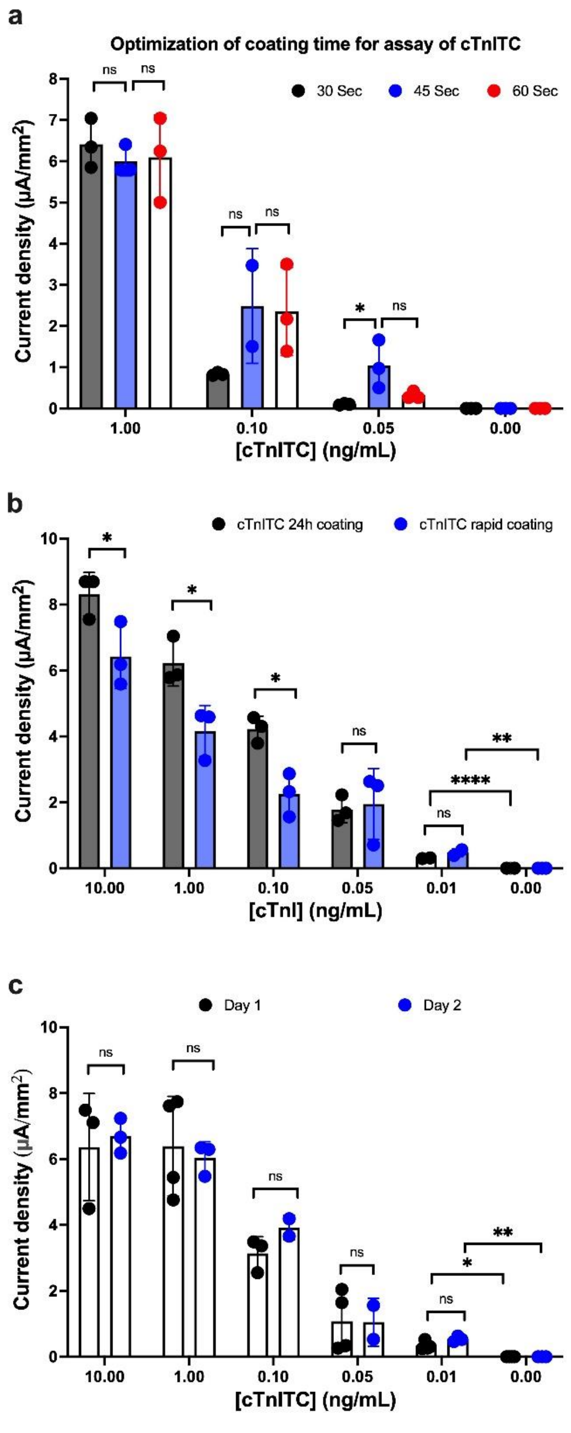


**Figure S5:** Characterization of coating and assay on EC biosensor. a) Optimization of coating time for sensors. Bar graph shows the assay of cTnITC with sensors coated with antifouling coating for 30 s (black dot), 45 s (blue dot), and 60 s (red dot). b) Comparison of rapid versus 24h coating for assay of cTnITC. Bars are the mean current density for cTnITC done on EC biosensor with 24h coating (black dots) and EC biosensor with rapid coating (blue dots). c)

Stability of precipitated TMB for detection of cTnITC. Bars graph shows the mean current density for cTnITC measured just after the assay, day 1 (black dot) and 1 day after the assay, day 2 (blue dot). Error bars represent the s.d. of the mean; n = 3. Analysis was done using 4-Parameter Logistic (4PL) curve fitting. Significant difference was determined by unpaired two-tailed t-test (ns $P > 0.05$; $^{*}P<0.05$; $^{**}P<0.01$; $^{***}P<0.001$; $^{****}P<0.0001$).

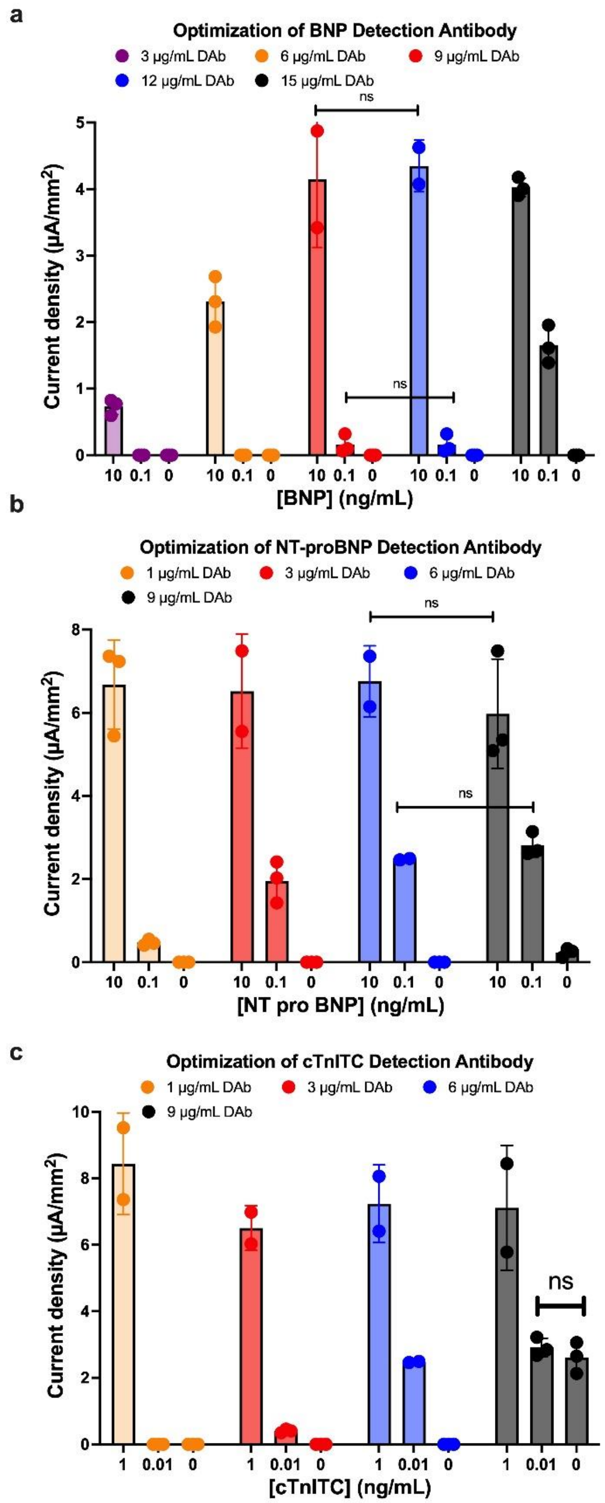


**Figure S6:** Optimization of assay condition for single step detection for different biomarkers in EC biosensor. a) Optimization of BNP detection antibody. Bar graph shows the mean current density for different concentration of detection antibody (3, 6, 9, 12, and 15 µg/mL) to perform assay of BNP at 3 different concentrations (10, 0.1, and 0 ng/mL). b) Optimization of NT-proBNP detection antibody. Bar graph shows the mean current density for different concentration of detection antibody (1, 3, 6, and 9 µg/mL) to perform assay of NT-proBNP at 3 different concentrations (10, 0.1, and 0 ng/mL). c) Optimization of cTnITC detection antibody. Bar graph shows the mean current density for different concentration of detection antibody (1, 3, 6, and 9 µg/mL) to perform assay of cTnITC at 3 different concentrations (1, 0.01, and 0 ng/mL). Error bars represent the s.d. of the mean; n=3. Significant difference was determined by unpaired two-tailed t-test (ns P > 0.05; *P<0.05; **P<0.01; ***P<0.001; ****P<0.0001).

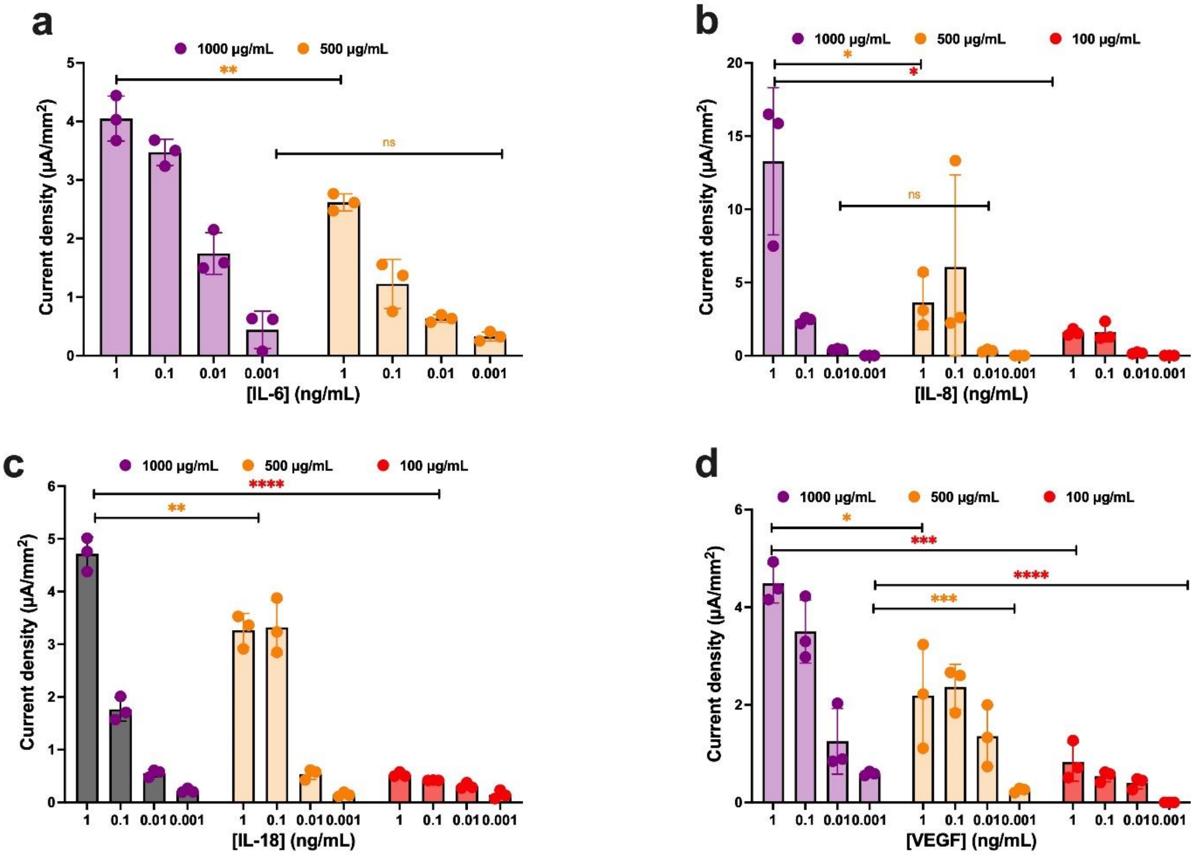


**Figure S7:** Optimization of capture antibody concentration for single step detection for different biomarkers in EC biosensor. a) Optimization of IL-6 capture antibody. Bar graph shows the mean current density for different concentration of IL-6 capture antibody (1000 and 500 µg/mL) to perform assay of Il-6 at 4 different concentrations (1, 0.1, 0.01, and 0.001 ng/mL). b) Optimization of IL-8 capture antibody. Bar graph shows the mean current density for different concentration of IL-8 capture antibody (1000, 500, and 100 µg/mL) to perform assay of IL-8 at 4 different concentrations (1, 0.1, 0.01, and 0.001 ng/mL). c) Optimization of IL-18 capture antibody. Bar graph shows the mean current density for different concentration of IL-18 capture antibody (1000, 500, and 100 µg/mL) to perform assay of IL-18 at 4 different concentrations (1, 0.1, 0.01, and 0.001 ng/mL). d) Optimization of VEGF capture antibody. Bar graph shows the mean current density for different concentration of VEGF capture antibody (1000, 500, and 100 µg/mL) to perform assay of VEGF at 4 different concentrations (1, 0.1, 0.01 and 0.001 ng/mL). Error bars represent the s.d. of the mean; n=3. Significant difference was determined by unpaired two-tailed t-test (ns P > 0.05; *P<0.05; **P<0.01; ***P<0.001; ****P<0.0001).

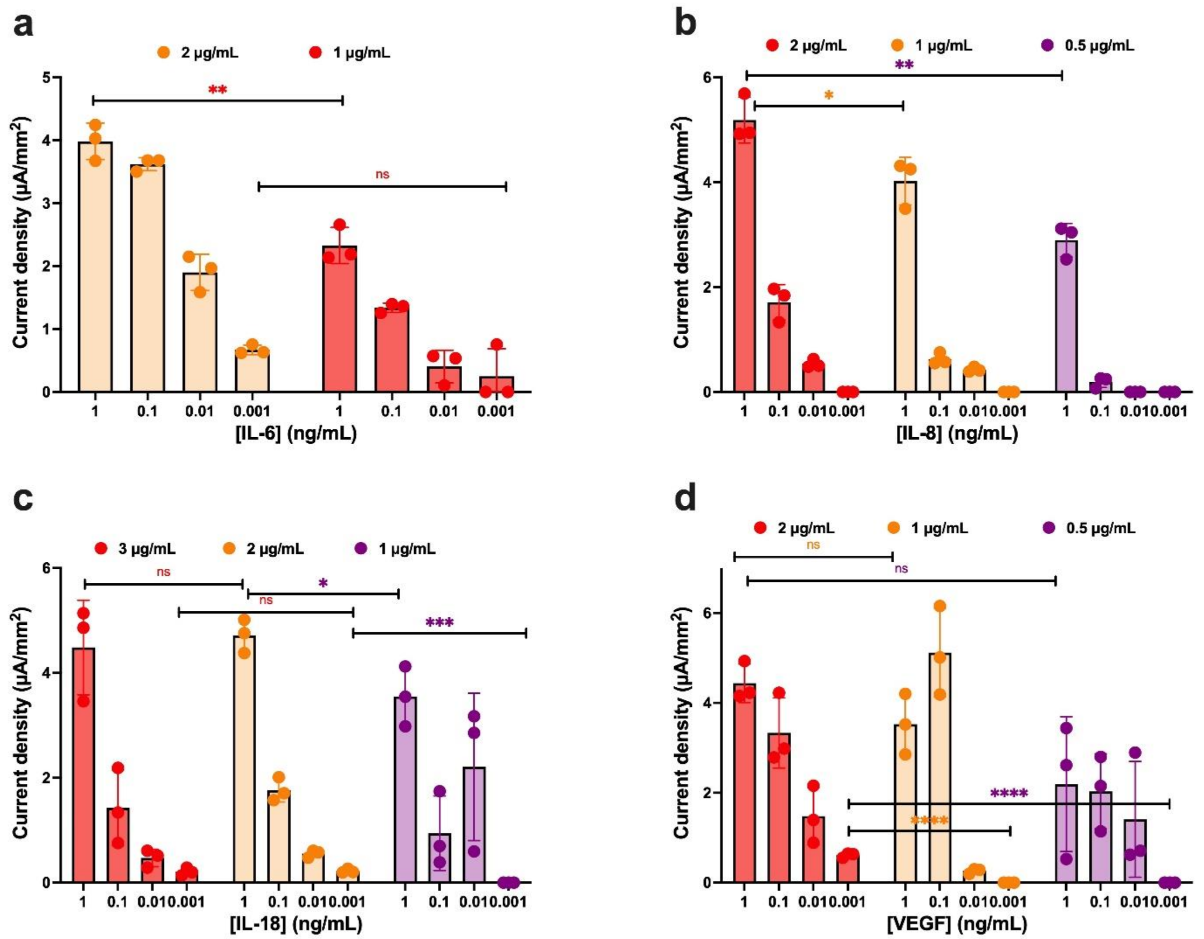


**Figure S8:** Optimization of detection antibody concentration for single step detection for different biomarkers in EC biosensor. a) Optimization of IL-6 detection antibody. Bar graph shows the mean current density for different concentration of IL-6 detection antibody (2 and 1 µg/mL) to perform assay of Il-6 at 4 different concentrations (1, 0.1, 0.01, and 0.001 ng/mL). b) Optimization of IL-8 detection antibody. Bar graph shows the mean current density for different concentration of IL-8 detection antibody (2, 1, and 0.5 µg/mL) to perform assay of IL-8 at 4 different concentrations (1, 0.1, 0.01, and 0.001 ng/mL). c) Optimization of IL-18 detection antibody. Bar graph shows the mean current density for different concentration of IL-18 detection antibody (3, 2, and 1 µg/mL) to perform assay of IL-18 at 4 different concentrations (1, 0.1, 0.01, and 0.001 ng/mL). d) Optimization of VEGF detection antibody. Bar graph shows the mean current density for different concentration of VEGF detection antibody (2, 1, and 0.5 µg/mL) to perform assay of VEGF at 4 different concentrations (1, 0.1, 0.01 and 0.001 ng/mL).

Error bars represent the s.d. of the mean; n=3. Significant difference was determined by unpaired two-tailed t-test (ns P > 0.05; *P<0.05; **P<0.01; ***P<0.001; ****P<0.0001).

Table 1: List of different biomarkers with their cut-off values and LOD obtained in EC Biosensor

| **Biomarkers** | **LOD of EC Biosensor (pg/mL)** | **Cut-off values** | **References** |
|---|---|---|---|
| Interleukin-6 | 3 | 1-40 pg/ml | [1] |
| Interleukin-8 | 9 | 1-40 pg/ml | [1a, 1c] |
| Interleukin-18 | 5 | 450-2200 | [1a, 2] |
| VEGF | 17 | 5-25 | [1a, 3] |
| PCT | 4 | 200-389 | [1b, 4] |
| GFAP | 2-16 | 22-230 | [5] |
| NF-L | 0.3-10 | 10-500 | [6] |

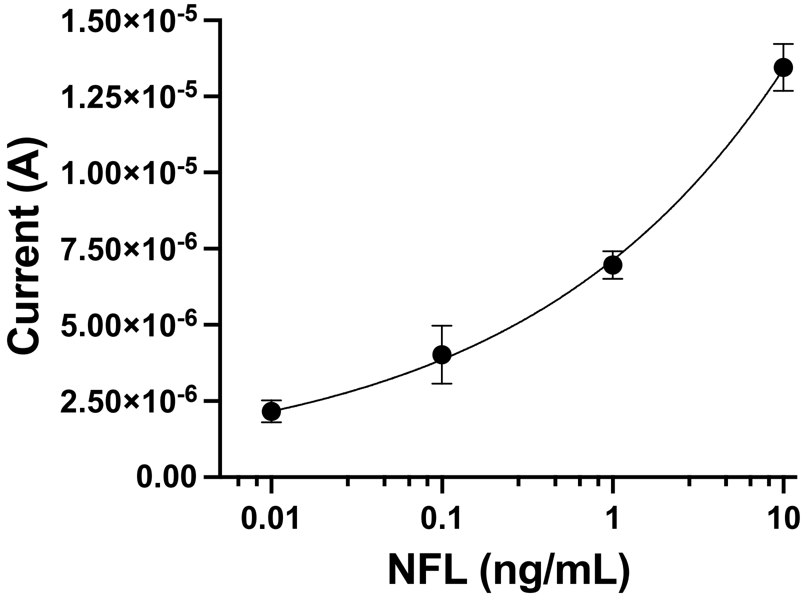


**Figure S9:** Calibration curves NF-L using antifouling nanocomposite coated Linxens Sensor. The left y-axis shows current intensity for different concentrations of NF-L run on coated sensor using unprocessed whole blood. Error bars represent the s.d. of the mean; n = 2. Analysis was done using 4-Parameter Logistic (4PL) curve fitting.

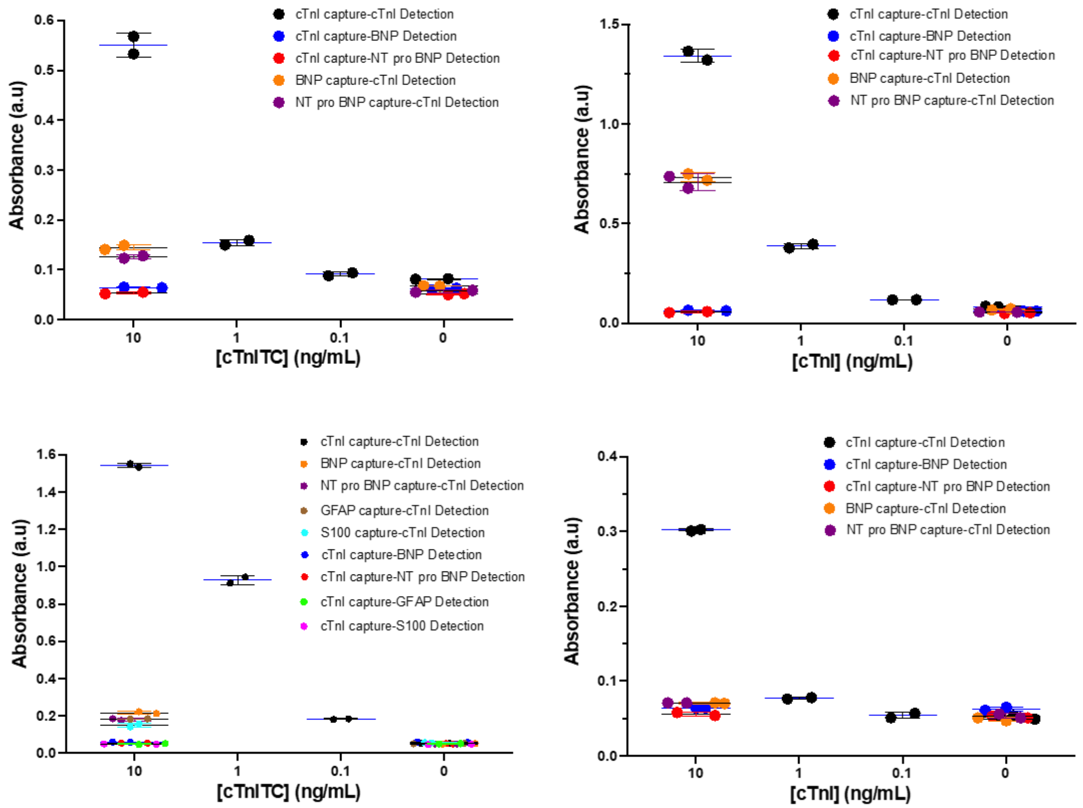


**Figure S10:** Specificity and cross-reactivity test for different Troponin antibody pair and antigen done in 96 well plate. a) Specificity and cross-reactivity of cTnITC antigen against different capture antibody (anti-NT-proBNP and anti-BNP) and detection antibody (anti-NT-proBNP and anti-BNP) along with specific detection with anti-cTnITC capture and detection antibody from abcam at different concentration of cTnITC. b) Specificity and cross-reactivity of cTnI antigen against different capture antibody (anti-NT-proBNP and anti-BNP) and detection antibody (anti-NT-proBNP and anti-BNP) along with specific detection with anti-cTnI capture and detection antibody from abcam at different concentrations of cTnI. c) Specificity and cross-reactivity of cTnITC antigen against different capture antibody (anti-NT-proBNP, anti-BNP, anti-GFAP, and anti-S-100) and detection antibody (anti-NT-proBNP, anti-BNP, anti-GFAP, and anti-S-100b) along with specific detection with anti-cTnITC capture and detection antibody from Advanced ImmunoChemical Inc at different concentration of cTnITC. d) Specificity and cross-reactivity of cTnI antigen against different capture antibody (anti-NT-proBNP and anti-BNP) and detection antibody (anti-NT-proBNP and anti-BNP) along with specific detection with anti-cTnI capture and detection antibody from Advanced ImmunoChemical Inc at different concentration of cTnI.

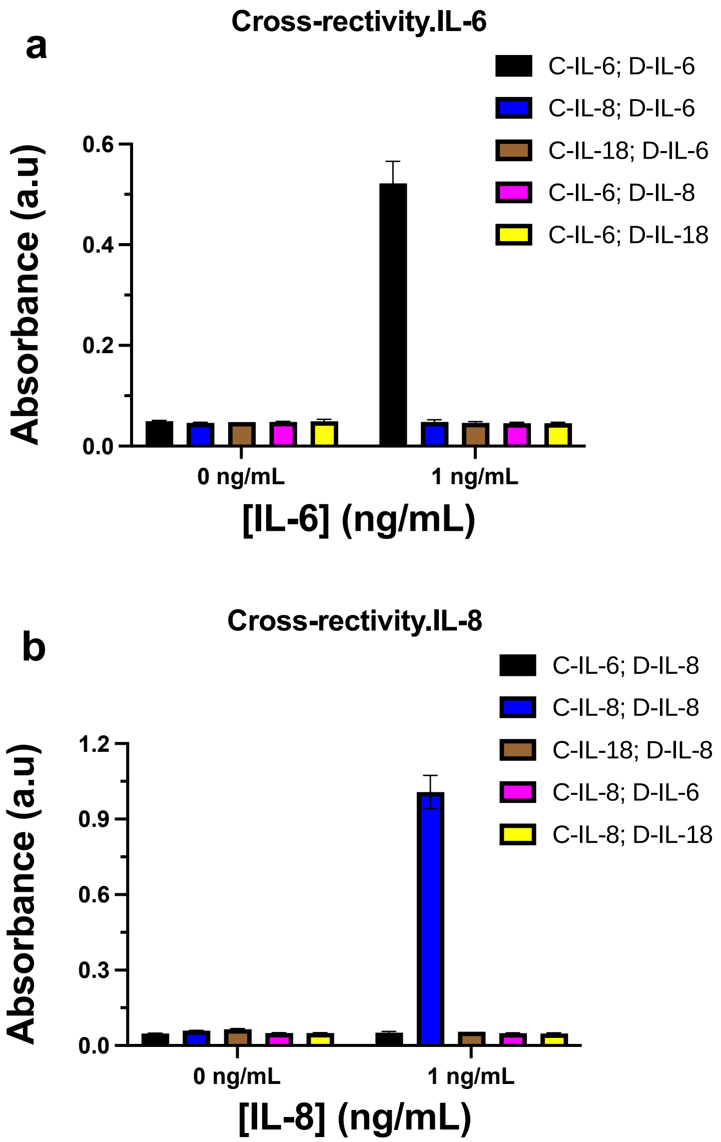


**Figure S11:** Specificity and cross-reactivity test for different Biomarkers of TB done in 96 well plate. a) Specificity and cross-reactivity of IL-6 antigen against different capture and detection antibodies (anti-IL-8 and anti-IL-18) along with specific detection with anti-IL-6 capture and detection antibody at different concentration of IL-6 (1 and 0 ng/mL). b) Specificity and cross-reactivity of IL-8 antigen against different capture and detection antibodies (anti-IL-6 and anti-IL-18) along with specific detection with anti-IL-8 capture and detection antibody at different concentration of IL-8 (1 and 0 ng/mL).

### References:


[1] a) R. Ahmad, L. Xie, M. Pyle, M. F. Suarez, T. Broger, D. Steinberg, S. M. Ame, M. G. Lucero, M. J. Szucs, M. MacMullan, *Science translational medicine* **2019**, 11, eaaw8287; b) N. I. Rashwan, M. H. Hassan, Z. M. M. El-Deen, A. El-Abd Ahmed, *Pediatrics & Neonatology* **2019**, 60, 149; c) A. S. Tanak, S. Muthukumar, S. Krishnan, K. L. Schully, D. V. Clark, S. Prasad, *Biosensors and Bioelectronics* **2021**, 171, 112726.

[2] a) M. Akgun, L. Saglam, H. Kaynar, A. K. Yildirim, A. Mirici, M. Gorguner, M. Meral, K. Ozden, *Respirology* **2005**, 10, 295; b) S. Wawrocki, M. Seweryn, G. Kielnierowski, W. Rudnicka, M. Druszczynska, *Saudi journal of biological sciences* **2020**, 27, 3035.

[3] a) C.-H. Lin, C.-C. Shu, C.-L. Hsu, S.-L. Cheng, J.-Y. Wang, C.-J. Yu, L.-N. Lee, *Scientific reports* **2016**, 6, 1; b) F. Alatas, M. Metintas, S. Erginel, H. Yildirim, *Chest* **2004**, 125, 2156.

[4] N. Kumar, R. Dayal, P. Singh, S. Pathak, V. Pooniya, A. Goyal, R. Kamal, K. Mohanty, *The Indian Journal of Pediatrics* **2019**, 86, 177.

[5] a) S. Çevik, M. M. Özgenç, A. Güneyk, Ş. Evran, E. Akkaya, F. Çalış, S. Katar, C. Soyalp, H. Hanımoğlu, M. Y. Kaynar, *Clinical neurology and neurosurgery* **2019**, 183, 105380; b) J. J. Bazarian, P. Biberthaler, R. D. Welch, L. M. Lewis, P. Barzo, V. Bogner-Flatz, P. G. Brolinson, A. Büki, J. Y. Chen, R. H. Christenson, *The Lancet Neurology* **2018**, 17, 782.

[6] a) A. Brodovitch, J. Boucraut, E. Delmont, A. Parlanti, A.-M. Grapperon, S. Attarian, A. Verschueren, *Scientific reports* **2021**, 11, 1; b) L. Gaetani, P. Eusebi, A. Mancini, L. Gentili, A. Borrelli, L. Parnetti, P. Calabresi, P. Sarchielli, K. Blennow, H. Zetterberg, *Multiple Sclerosis and Related Disorders* **2019**, 35, 228; c) S. Thebault, M. Abdoli, S.-M. Fereshtehnejad, D. Tessier, V. Tabard-Cossa, M. S. Freedman, *Scientific Reports* **2020**, 10, 1.
